# Causal influence of dietary habits on the risk of major depressive disorder: A diet-wide Mendelian randomization analysis

**DOI:** 10.1101/2020.07.12.20150367

**Authors:** Tzu-Ting Chen, Chia-Yen Chen, Chiu-Ping Fang, Ying-Chih Cheng, Yen-Feng Lin

## Abstract

**Background & aims:** Some evidence suggests that diet may potentially increase or decrease the risk of major depressive disorder (MDD). However, the association between dietary habits and MDD remains controversial. The aim of this study is to systemically investigate the causal influence of dietary habits on the risk of MDD by Mendelian randomization (MR) using diet- and genome-wide summary data.

**Methods:** To perform two-sample MR, we collected publicly available genome-wide association studies’ summary statistics for dietary habits from Benjamin Neale’s lab (*n* = 361,194) and MDD from the Psychiatric Genomics Consortium *(n =* 142,646). We used a weighted median approach to synthesize MR estimates across genetic instruments. For the robustness of our results, we compared weighted median results with results from the inverse-variance weighted method, the weighted mode method, and MR-PRESSO.

**Results:** Beef intake showed a significant protective effect against MDD (β = -1.25; p-value = 0.002; Bonferroni-corrected p-value = 0.034; 9 single nucleotide polymorphisms [SNPs]); and cereal intake was nominally significantly protective (β = -0.52; p-value = 0.011; 21 SNPs). In contrast, non-oily fish intake showed a nominally significantly effect on the risk of MDD (β = 0.84; p-value = 0.030; 6 SNPs). We obtained similar results by using an inverse-variance weighted method and weighted mode approach, although some results were non-significant. On the other hand, we did not observe any significant causal effect of MDD on dietary habits.

**Conclusions:** In this two-sample MR analysis, we observed that higher beef and cereal intake may be protective factors for MDD, and that higher non-oily fish intake might increase the risk for MDD. However, MDD did not appear to affect dietary habits. Potential mechanisms need to be further investigated to support our novel findings.

## 1. Introduction

Globally, major depressive disorder (MDD) has been one of the leading causes of non-fatal health loss for nearly three decades [1]. In 2019, the prevalent cases and incident cases of MDD were approximately 185.2 million and 274.8 million, respectively. The number of all-age years lived with disability attributed to MDD increased by 15.6% from 2010 to 2019 [1]. Considering issues related to current treatments for depression, such as adverse side effects and unsatisfactory response rates to antidepressant medication, many studies have suggested that non-pharmacological prevention strategies, such as physical activity and healthy diet, may play an important role in reducing the disease burden of MDD.

In the past decade, a number of nutritional epidemiological studies have suggested that diet may potentially increase or decrease the risk of MDD. It is hypothesized that nutrition may activate hormonal, neurotransmitter, and signaling pathways in the gut and then modulate depression-associated biomarkers [2]. However, the findings are inconsistent and inconclusive for many diets’ effects on MDD. For example, a meta-analysis of observational studies reported that increased meat consumption was associated with higher incidence but not prevalence of depression [3]. Meanwhile, an observational study in females reported that consumption of less red meat than the recommended intake may be associated with an increased risk of depression [4]. Finally, a recent review of the association between meat abstention and depression showed mixed results; however, the majority of studies, and especially the most rigorous ones, demonstrated a higher risk of depression in those who avoided meat [5]. Similarly, alcohol consumption was shown to be associated with increased risk of MDD in some studies [6, 7, 8, 9] but not in the others [10, 11, 12]. Several other potential protective factors for MDD were identified through randomized controlled trials and observational studies, including coffee and caffeine intake [13], omega-3 polyunsaturated fatty acid intake [14], fruit and vegetable intake [15], frequent fish consumption [16], plain water intake [17], and decreased fat intake (with induced body weight loss) [18]. However, there were also studies showing null association of MDD with consumption of tea and coffee [19], as well as fish [20].

In nutritional epidemiology studies, minimizing confounding bias that may contribute to inconsistent results regarding the relationship between diet and MDD is very challenging. Recently, several genome-wide association studies (GWAS) have indicated that dietary habits are heritable traits [21, 22]; therefore, Mendelian randomization (MR), which leverages genetic instruments to reduce potential confounding biases, may be an appropriate study design to evaluate the effects of diet on a disease or a health outcome [23]. The aim of this study is to systemically investigate the causal influence of dietary habits on the risk of MDD by the MR approach using diet- and genome-wide summary data.

## 2. Materials and methods

### 2.1. Study design

We applied a two-sample MR study design that uses genetic variants as instrumental variables (IVs) to investigate the causal relationship between exposure and outcome [24, 25, 26]. We leveraged summary statistics from large-scale GWAS meta-analyses to increase statistical power in the two-sample MR study. Our MR method is described in detail in the Supplemental Methods.

### 2.2. Dietary habits GWAS summary statistics

We obtained genome-wide associations for 20 dietary habits from Benjamin Neale’s lab (http://www.nealelab.is/uk-biobank/), and the questions used to assess these dietary habits were summarized in Supplementary Table S1. The lab included GWAS results from the UK Biobank, using data of 361,194 participants of European ancestry, with 13.7 million QC-passing significant single nucleotide polymorphisms (SNPs) (see also the Supplemental Methods). We further filtered the summary statistics to include only SNPs with minor allele frequency ≥ 0.01. We selected genome-wide significant SNPs with association p-value < 5 × 10^-8^.

### 2.3. Major depressive disorder GWAS summary statistics

The Psychiatric Genomics Consortium (PGC) provided summary statistics from a genome-wide association meta-analysis of MDD (see the Supplementary Methods for cohort details) [27]. This meta-analysis included 45,396 MDD patients and 97,250 controls of European ancestry, with 9.8 million genetic variants. We filtered the data to include only imputed and genotyped variants with minor allele frequency ≥ 0.01 and imputation information score ≥ 0.8. We used a relaxed threshold of association p-value < 1×10^-6^, which has been used in previous MR studies to select genetic instruments when few significant loci are available [28, 29, 30].

### 2.4. Statistical analysis

First, we performed linkage disequilibrium score regression (LDSC) [31, 32] with the default settings on the GWAS summary data of dietary habits and MDD to estimate the SNP-based heritability of each trait and genetic correlations between each dietary habit and MDD. Next, we assessed the bidirectional relationships between dietary habits and MDD by two-sample MR. For the MR analysis, we performed linkage disequilibrium clumping on significant SNPs to keep independent SNPs as genetic instruments. When the SNPs were not available in the outcome summary data, we replaced them with proxy SNPs in the highest linkage disequilibrium with r^2^ ≥ 0.8. We harmonized exposure and outcome data, and aligned the effect allele in exposure and outcome GWAS. We inferred positive strand alleles using allele frequencies for palindromic SNPs, and removed palindromic SNPs with a minor allele frequency > 0.42, given that such alleles cannot be reliably aligned. A weighted median approach was used to synthesize MR estimates. For the robustness of our results, we compared weighted median results with other estimates, including an inverse-variance weighted (IVW) method and a weighted mode method.

In an MR study, it is essential to check for horizontal pleiotropy, which will lead to biased estimates. We used a MR-PRESSO test to detect horizontal pleiotropic outliers in the multi-instrument MR study and to correct for horizontal pleiotropy via outlier removal [33]. In addition, we examined horizontal pleiotropy by test for the intercept in the MR-Egger regression, MR-Egger test and Cochran’s Q test for heterogeneity, and the leave-one-out analysis [34]. All analyses were performed using the statistical software R with the TwoSampleMR package for processing and harmonizing exposure and outcome data, and conducting MR analyses with the ieugwasr package for linkage disequilibrium clumping.

## 3. Results

The estimated SNP-based heritability of dietary habits ranged from 0.025 to 0.080 (see Table 1). Significant genetic correlation with MDD was found for fresh fruit intake (rg = -0.130; p-value = 0.001; Bonferroni-corrected p-value = 0.016), dried fruit intake (rg = - 0.138; p-value < 0.001; Bonferroni-corrected p-value = 0.006), lamb/mutton intake (rg = - 0.162; p-value = 0.001; Bonferroni-corrected p-value = 0.012), cheese intake (rg = -0.130; p-value < 0.001; Bonferroni-corrected p-value = 0.002), cereal intake (rg = -0.193; p-value < 0.001; Bonferroni-corrected p-value < 0.001), salt added to food (rg = 0.142; p-value < 0.001; Bonferroni-corrected p-value = 0.006), hot drink temperature (rg = 0.098; p-value = 0.002; Bonferroni-corrected p-value = 0.042), and alcohol intake frequency (rg = 0.235; p-value < 0.001; Bonferroni-corrected p-value < 0.001) (see Table 1).

**Table 1.** The estimated SNP-based heritability of dietary habits and genetic correlation with major depressive disorder.

| **Dietary habits** | **Heritability** | **Genetic correlation** | **p-value** |
| --- | --- | --- | --- |
| Cooked vegetable intake | 0.038 (0.002) | 0.112 (0.045) | 0.013 |
| Salad / raw vegetable intake | 0.044 (0.002) | -0.089 (0.045) | 0.050 |
| Fresh fruit intake | 0.060 (0.003) | -0.130 (0.039) | 0.001 |
| Dried fruit intake | 0.063 (0.003) | -0.138 (0.038) | <0.001 |
| Oily fish intake | 0.063 (0.003) | -0.060 (0.037) | 0.111 |
| Non-oily fish intake | 0.025 (0.002) | 0.034 (0.052) | 0.522 |
| Processed meat intake | 0.043 (0.003) | -0.012 (0.041) | 0.775 |
| Poultry intake | 0.032 (0.002) | -0.031 (0.051) | 0.545 |
| Beef intake | 0.036 (0.002) | -0.040 (0.052) | 0.445 |
| Lamb/mutton intake | 0.041 (0.002) | -0.162 (0.047) | 0.001 |
| Pork intake | 0.029 (0.002) | -0.082 (0.055) | 0.132 |
| Cheese intake | 0.067 (0.003) | -0.130 (0.033) | <0.001 |
| Bread intake | 0.049 (0.003) | 0.049 (0.041) | 0.231 |
| Cereal intake | 0.059 (0.003) | -0.193 (0.038) | <0.001 |
| Salt added to food | 0.080 (0.004) | 0.142 (0.040) | <0.001 |
| Tea intake | 0.057 (0.003) | 0.081 (0.037) | 0.028 |
| Coffee intake | 0.048 (0.003) | -0.077 (0.040) | 0.054 |
| Hot drink temperature | 0.076 (0.003) | 0.098 (0.032) | 0.002 |
| Water intake | 0.062 (0.003) | 0.050 (0.036) | 0.169 |
| Alcohol intake frequency | 0.080 (0.004) | 0.235 (0.036) | <0.001 |

<sup>a</sup> Bonferroni-corrected p-value for genetic correlations between dietary habits and major depressive disorder: fresh fruit intake is 0.016; dried fruit intake is 0.006; lamb/mutton intake is 0.012; cheese intake is 0.002; cereal intake is < 0.001; salt added to food is 0.006; hot drink temperature is 0.042; alcohol intake frequency is < 0.001.

The weighted median MR analysis showed a causally protective effect of beef intake on MDD after Bonferroni correction (β = -1.25; p-value = 0.002; Bonferroni-corrected p-value = 0.034; 9 SNPs) (Table 2). We also found cereal intake showing a protective effect on MDD, with a nominally significant p-value (β = -0.52; p-value = 0.011; 21 SNPs) (Table 2). In contrast, non-oily fish intake showed a nominally significantly effect on the risk of MDD (β = 0.84; p-value = 0.030; 6 SNPs). The causal effect estimates on MDD from each SNP and the distribution of genetic effects on exposure and outcome are summarized in Figures 1, Supplementary Figures S1 and S2 for beef intake, non-oily fish intake, and cereal intake, respectively. We obtained similar results by using the IVW method and weighted mode approach, despite some results being non-significant. SNPs selected as instrumental variables for MR analyses for effects of dietary habits on MDD are listed in Supplementary Tables S2– S21.

**Table 2.** Mendelian randomization results by using the weighted median approach for estimating the causal effect of dietary habits on major depressive disorder.

| **Dietary habits** | **Number of SNPs** | **$\beta$** | **Standard error** | **p-value** |
| --- | --- | --- | --- | --- |
| Cooked vegetable intake | 6 | 0.227 | 0.517 | 0.660 |
| Salad / raw vegetable intake | 11 | 0.672 | 0.401 | 0.094 |
| Fresh fruit intake | 29 | 0.179 | 0.288 | 0.534 |
| Dried fruit intake | 27 | -0.269 | 0.245 | 0.272 |
| Oily fish intake | 33 | -0.051 | 0.157 | 0.744 |
| Non-oily fish intake | 6 | 0.837 | 0.385 | 0.030 |
| Processed meat intake | 8 | -0.048 | 0.316 | 0.880 |
| Poultry intake | 3 | 0.776 | 0.590 | 0.188 |
| Beef intake<sup>a</sup> | 9 | -1.252 | 0.399 | 0.002 |
| Lamb/mutton intake | 14 | 0.562 | 0.397 | 0.156 |
| Pork intake | 5 | 0.382 | 0.643 | 0.553 |
| Cheese intake | 30 | -0.305 | 0.165 | 0.065 |
| Bread intake | 16 | -0.235 | 0.251 | 0.348 |
| Cereal intake | 21 | -0.517 | 0.204 | 0.011 |
| Salt added to food | 61 | 0.146 | 0.129 | 0.255 |
| Tea intake | 23 | -0.059 | 0.142 | 0.676 |
| Coffee intake | 25 | 0.025 | 0.168 | 0.882 |
| Hot drink temperature | 39 | -0.054 | 0.259 | 0.834 |
| Water intake | 24 | 0.038 | 0.180 | 0.832 |
| Alcohol intake frequency | 52 | -0.002 | 0.077 | 0.982 |

SNP, single nucleotide polymorphism
<sup>a</sup> The Bonferroni-corrected p-value for the effect of beef intake on major depressive disorder is 0.034.

**Figure 1.**
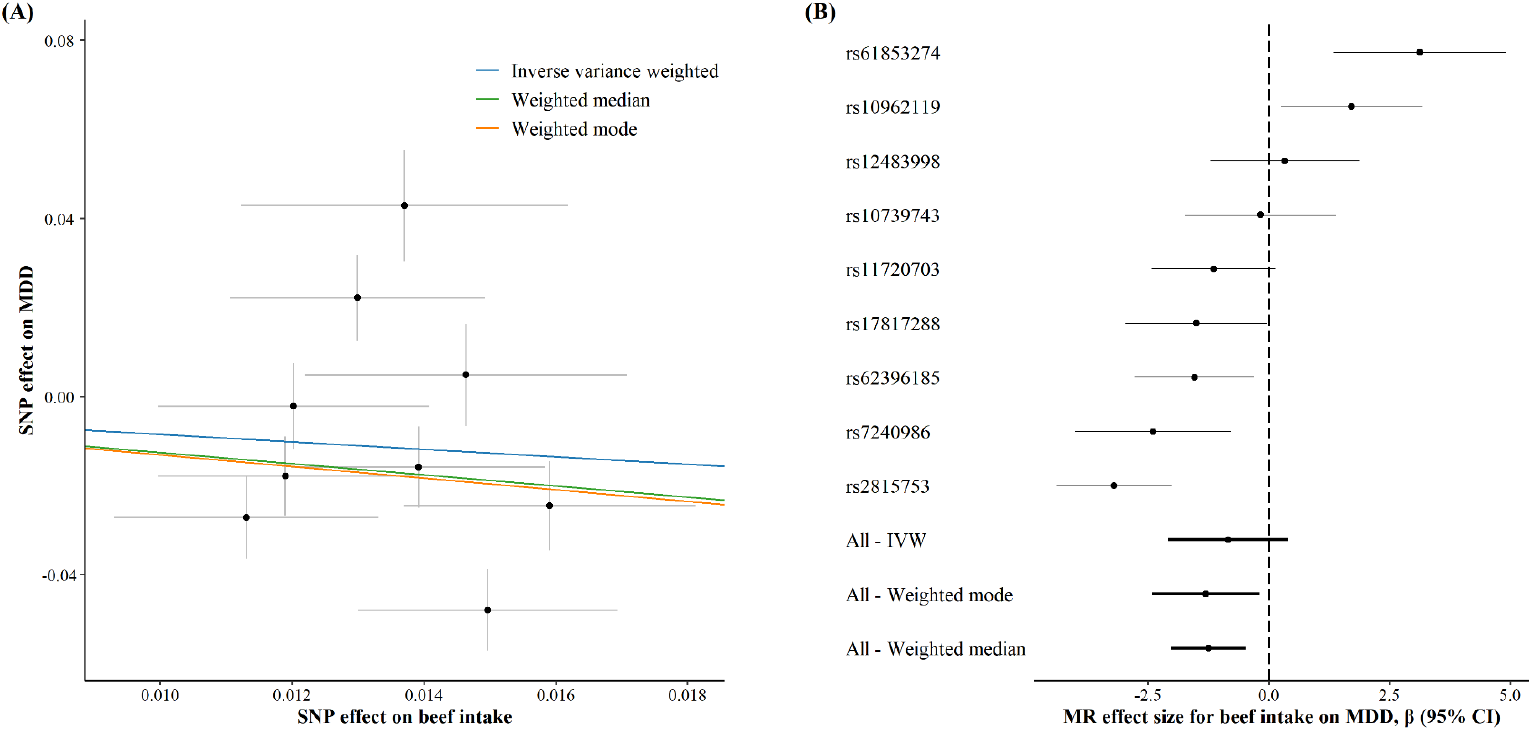
Mendelian randomization results for estimating the causal effects of beef intake on major depressive disorder. (A) Scatter plot showing the effects of SNPs on beef intake versus major depressive disorder. (B) Forest plot of Mendelian randomization effect size for beef intake on major depressive disorder

We also assessed the effects of MDD as an exposure on these 20 dietary habits (Table 3). We did not observe a significant effect of MDD on any of these 20 dietary habits, and we obtained similar results by using the IVW method and weighted mode approach. SNPs selected as potential instrumental variables for MR analyses for effects of MDD on dietary habits are listed in Supplementary Table S22.

**Table 3.** Mendelian randomization results by using the weighted median approach for estimating the causal effect of major depressive disorder on dietary habits.

| **Dietary habits** | **$\beta^a$** | **Standard error** | **p-value** |
| --- | --- | --- | --- |
| Cooked vegetable intake | 0.020 | 0.013 | 0.119 |
| Salad / raw vegetable intake | -0.004 | 0.013 | 0.759 |
| Fresh fruit intake | 0.006 | 0.011 | 0.547 |
| Dried fruit intake | -0.013 | 0.014 | 0.349 |
| Oily fish intake | 0.001 | 0.019 | 0.977 |
| Non-oily fish intake | 0.019 | 0.014 | 0.179 |
| Processed meat intake | 0.035 | 0.019 | 0.061 |
| Poultry intake | -0.008 | 0.015 | 0.606 |
| Beef intake | 0.001 | 0.015 | 0.958 |
| Lamb/mutton intake | -0.022 | 0.012 | 0.076 |
| Pork intake | -0.020 | 0.013 | 0.120 |
| Cheese intake | -0.031 | 0.017 | 0.065 |
| Bread intake | 0.009 | 0.018 | 0.600 |
| Cereal intake | 0.006 | 0.014 | 0.661 |
| Salt added to food | 0.007 | 0.019 | 0.709 |
| Tea intake | 0.021 | 0.021 | 0.311 |
| Coffee intake | 0.003 | 0.013 | 0.832 |
| Hot drink temperature | 0.004 | 0.011 | 0.681 |
| Water intake | 0.006 | 0.016 | 0.700 |
| Alcohol intake frequency | 0.032 | 0.028 | 0.250 |

<sup>a</sup> Included 18 single nucleotide polymorphisms meeting the relaxed threshold (p-value < 1 ×

To detect horizontal pleiotropy, we employed the MR-PRESSO global test, which detected three potential outliers in the MR effect of beef intake on MDD (Supplementary Table S23). The effect estimates for beef intake on MDD (β = -1.31; p-value = 0.001) remained significant after correcting for horizontal pleiotropy via outlier removal. The MR-PRESSO distortion test showed that the distortion in the causal estimates before and after outlier removal was not significant. Moreover, we examined the horizontal pleiotropy by using an MR-Egger intercept test, which showed that the horizontal pleiotropy is negligible. We investigated the horizontal pleiotropy by Cochran’s Q statistic, the MR-Egger test approach (Supplementary Table S23), and funnel plots (Supplementary Figures S3–S5). The results also show that horizontal pleiotropy may not exist on these effects after correcting for horizontal pleiotropy via outlier removal.

In our sensitivity analysis, we conducted leave-one-out analyses, excluding one significant SNP each time. Although a few MR leave-one-out estimates were not statistically significant, we observed a similar trend for the effects of beef intake on MDD (Figure 2) and non-oily fish intake (Supplementary Figure S6) across them. The effects of cereal intake on MDD (Supplementary Figure S7) in leave-one-out analyses were similar to the main MR results.

**Figure 2.**
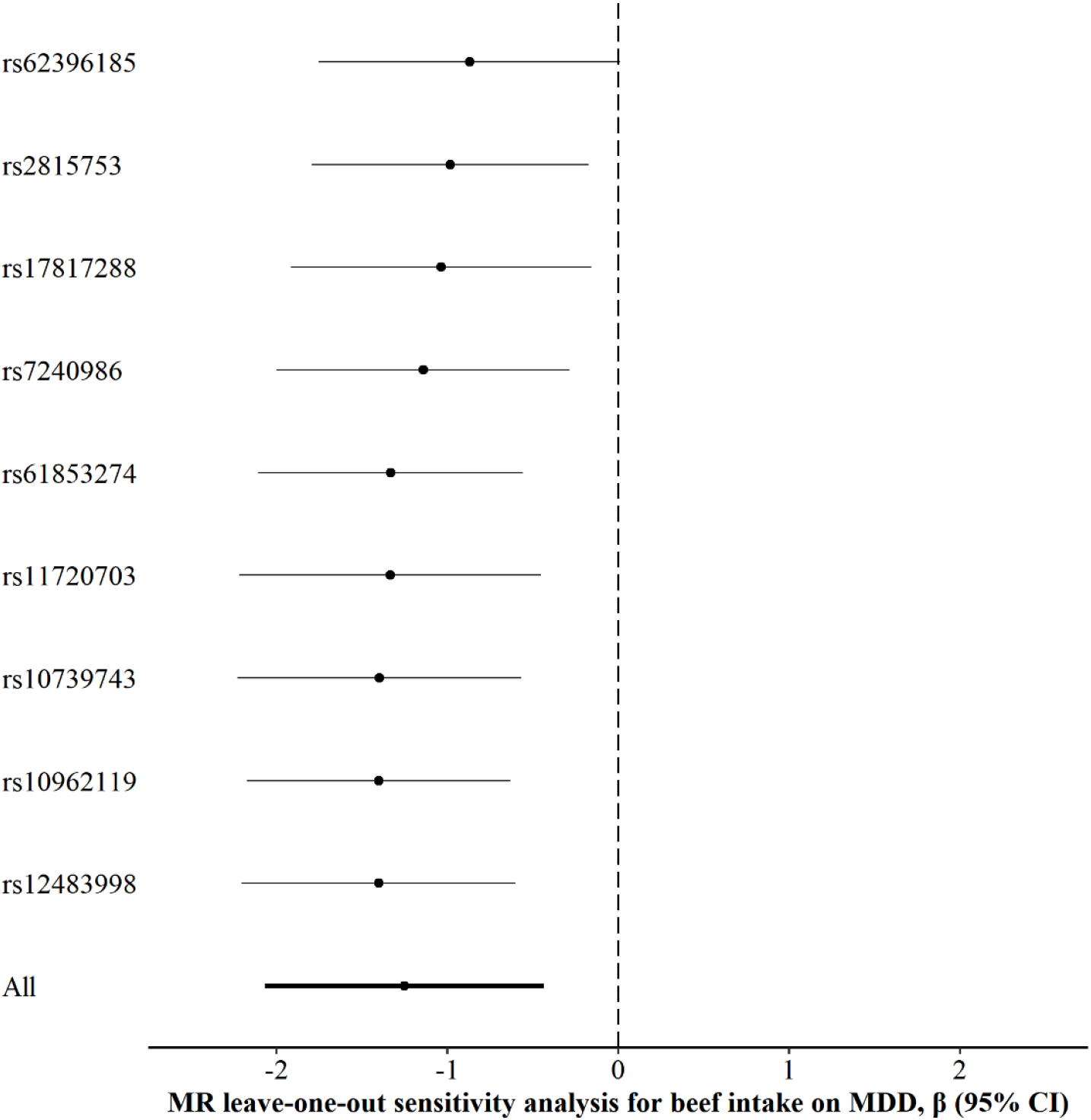
Leave-one-out analysis of the effect of beef intake on major depressive disorder.

## 4. Discussion

In this two-sample MR using the largest diet GWAS results available to date, we systemically examined the causal relationships between 20 dietary habits and MDD. We observed potential protective effects of beef intake and cereal intake on MDD. We also observed the effect of non-oily fish intake on the risk of MDD, while other dietary habits did not show significant effects on MDD. The results were consistent after we corrected for horizontal pleiotropy via outlier removal by using MR-PRESSO. On the other hand, we did not observe that MDD had a significant effect on any of those 20 dietary habits. In addition, a higher risk of MDD was found to be genetically correlated with higher hot drink temperature, more intake of salt added to food, and more intake of alcohol, as well as less intake of fresh fruit, dried fruit, lamb/mutton, cheese, and cereal. However, these genetic correlations do not necessarily indicate a causal relationship but are more likely attributable to pleiotropy.

We found a significant protective effect of beef intake on MDD, but consumptions of processed meat, poultry, lamb/mutton, and pork were not seen to significantly influence MDD. Previous studies show inconsistent results for the effect of meat intake on MDD, which may be partly due to differences in study designs and methods, including different food frequency questionnaires, diagnostic tools for depression, and different residual confounding [3, 4, 35, 36]. In the present study, residual confounding was minimized by applying a two-sample MR study design that used genetic variants as IVs for meat intake to estimate the causal relationship between the consumption of each category of meat and MDD. In addition, meat consumption in the current study was divided into five different exposures, each for a different type of meat. This distinction of meat type was not always considered in previous studies and may contribute to the inconsistent results among studies. There are several potential mechanisms that may possibly explain the observed beneficial effect of beef consumption on MDD. Beef contains nutrients that may be beneficial in the prevention of MDD, such as zinc, iron, and protein. A meta-analysis found that dietary zinc and iron intake significantly decrease the risk of depression [37]. In addition, sufficient protein intake is essential to building and maintaining lean muscle mass, while lower lean muscle mass has been reported to be associated with more depressive symptoms [38]. To the best of our knowledge, this is the first study reporting a potential beneficial causal effect of beef intake on lowering the risk of MDD. Therefore, our MR results need to be interpreted cautiously, especially considering the potential pleiotropic bias in MR analysis. Further investigation of the causal relationship between beef intake and MDD is also warranted in order to validate the results.

In the present study, we found that cereal intake might decrease the risk of MDD, with a nominally significant p-value. Previous studies show inconclusive evidence for the effect of cereal intake on MDD. Although it has been suggested that increasing consumption of wholegrain cereals may prevent depression [39], several other studies have shown either a null effect or protective effect of cereal intake on depression only in older adults [40, 41, 42]. One possible explanation for the potential protective effect is that cereal fiber may modulate gut microbiota [43] and subsequently influence depressive illness through the mediation of gut microbiota [44]. This hypothesis needs to be confirmed in further studies.

Moreover, we found that non-oily fish might increase the risk of MDD, with a nominally significant p-value. To our knowledge, no previous studies were aimed at evaluating the effect of non-oily fish intake on MDD, and further studies are needed to validate our findings and gain mechanistic insights. In addition, our MR analysis showed no significant effect of oily fish on MDD. Omega-3 polyunsaturated fatty acids, especially adequate eicosapentaenoic acid (EPA), are generally suggested as being beneficial against depression [14]. However, for dietary fish intake, a randomized controlled trial showed that oily fish consumption is not associated with the risk of MDD [20], and a dose–response meta-analysis of 10 prospective cohort studies showed an increment of one serving per week of fish a non-significant effect on depression [45]. These negative findings are consistent with our MR results and may be due to insufficient dosage of EPA from dietary fish consumption.

Some evidence suggests that fruit, vegetable, coffee, water, and moderate alcohol consumption are protective factors for MDD [10, 11, 12, 13, 15, 17]; but several other studies have failed to replicate these associations [6, 7, 8, 9, 19]. Considering potential residual bias, insufficient power, and a lack of temporality in cross-sectional studies, a carefully designed study with a large sample size, such as an MR study, is therefore required to help resolve this controversial issue.

Our study may suffer from the common limitations of MR studies; that is, assumptions may be violated due to horizontal pleiotropy. A genetic variant affecting the outcome through a different pathway from the exposure under investigation could lead to biased estimates. Consequently, we did several sensitivity analyses to detect and minimize the bias, including a MR-PRESSO test, an intercept test in the MR-Egger regression, a heterogeneity test, and a leave-one-out analysis; these produced consistent results. Second, while the effects of diet on MDD may vary across sex and age groups, we examined only the overall effects adjusted for sex and age due to limitation of publicly available GWAS summary statistics. Furthermore, the present study included only individuals of European ancestry; thus, the results might lack generalizability to other populations. Last, as information about dietary habits was collected retrospectively by a shortened food frequency questionnaire, recall bias cannot be excluded. In addition, several dietary habits, depending on overall daily or weekly frequency, were regarded as ordinal variables, and the effect of SNPs on dietary habits was assumed to be linear. This strong assumption may bias the GWAS results toward the null and consequently result in a false negative result for the relationship between dietary habits and MDD. Thus, we suggest further studies to collect detailed dietary data prospectively and to test for non-linear associations between dietary habits and MDD.

Despite these limitations, to our knowledge this study is the most comprehensive MR study to date to evaluate the causal role of dietary habits on the risk of MDD. We leveraged summary statistics from large-scale GWAS meta-analysis to increase statistical power. In addition, sensitivity analyses found no substantial difference in the results from those of the main analysis, thereby indicating that our findings are robust. Identification of protective dietary habits for MDD is crucial for primary prevention; however, interpretation of the evidence of causality from our study needs to be done so cautiously. We have stressed the need for further investigation to confirm and generalize our findings.

## 5. Conclusion

In this two-sample MR analysis, we observed that higher intake of beef and cereal may be protective against MDD, and that higher non-oily fish intake might increase the risk for MDD. However, MDD does not appear to affect dietary habits. Further validation of these novel findings and investigations of potential mechanisms are required.

## Data Availability

We collected publicly available GWAS summary statistics for 20 dietary habits in UK Biobank from the website of Benjamin Neale's lab (http://www.nealelab.is/uk-biobank/). The PGC provided summary statistics from a genome-wide association meta-analysis, excluding 23andMe and UK Biobank, of MDD at https://www.med.unc.edu/pgc/download-results/.

## Acknowledgments

We would like to thank the National Core Facility for Biopharmaceuticals (NCFB, MOST 106-2319-B-492-002) and the National Center for High-performance Computing (NCHC) of the National Applied Research Laboratories (NARLabs) of Taiwan for providing computational resources and storage resources.

## Author contributions

**Tzu-Ting Chen:** Data curation, Formal analysis, Methodology, Software, Validation, Visualization, Writing (original draft preparation). **Chia-Yen Chen:** Conceptualization, Methodology, Validation, Writing (reviewing and editing). **Chiu-Ping Fang:** Data curation, Formal analysis, Visualization, Writing (original draft preparation). **Ying-Chih Cheng:** Conceptualization, Validation, Writing (reviewing and editing). **Yen-Feng Lin:** Conceptualization, Methodology, Validation, Visualization, Supervision, Writing (reviewing and editing). All authors gave final approval and agree to be accountable for all aspects of the work.

## Data availability

**Supplementary data to this article can be found online**.

