## supplemental material for "Causal influence of dietary habits on the risk of major depressive disorder: A diet-wide Mendelian randomization analysis"

**Supplemental Methods**

*Mendelian randomization*

A Mendelian randomization (MR) study design uses genetic variants as instrumental variables (IVs) for exposure to investigate the causal relationship between exposure and outcome. IVs are commonly used to minimize the potential confounding bias in observational studies. The following directed acyclic graph presents the causal relationship between genetic variants (G), exposure (E), outcome (Y), and confounding factors (U).

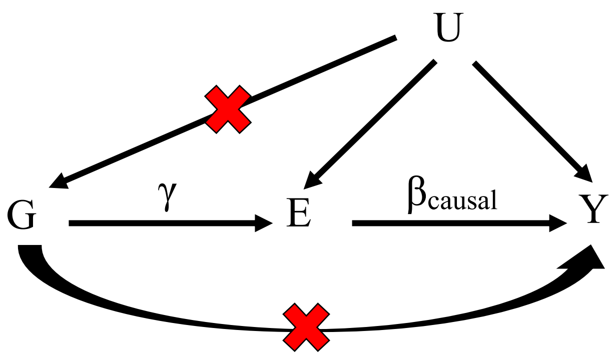


To investigate the effects of E on Y ($\beta_{causal}$), adjusting for U to obtain an unbiased estimate for $\beta_{causal}$ is needed in observation studies. However, it is difficult to directly adjust either unmeasured or unknown confounding factors. MR may be a solution to this issue, using the effect of G on E and the effect of G on Y to estimate the average effect of E on Y.

There are three essential definitions for an IV. First, G has a causal effect on E ($\gamma$). Next, G affects Y only through E; that is, there is neither a direct nor an effect of G on Y through other components. Third, G does not share common causes with Y; specifically, there is no direct effect of U on G. As a result, genetic variants would be suitable to be instrumental variables (IVs) in genetic studies if there are no pleiotropic effects.

*Dietary habits*

We obtained the results of genome-wide associations for 20 dietary habits — including cooked vegetable intake, salad or raw vegetable intake, fresh fruit intake, dried fruit intake, oily fish intake, non-oily fish intake, processed meat intake, poultry intake, beef intake, lamb or mutton intake, pork intake, cheese intake, bread intake, cereal intake, salt added to food, tea intake, coffee intake, hot drink temperature, water intake, and alcohol intake frequency — from Benjamin Neale's lab at Massachusetts General Hospital and the Stanley Center for Psychiatric Research, Broad Institute of MIT and Harvard (http://www.nealelab.is/uk-biobank/). The lab's website includes the results of genome-wide association studies (GWAS) from the UK Biobank, using data of 361,194 participants of

European ancestry with 13.7 million QC-passing single nucleotide polymorphisms (SNPs). In the UK Biobank, information on the 20 dietary habits was collected retrospectively by a shortened food frequency touchscreen questionnaire at baseline. Data on the 20 dietary habits consist of quantitative continuous variables, such as, on average, the number of heaped tablespoons of cooked vegetables per day, and ordinal non-quantitative variables depending on frequency, such as how often one eats oily fish. If the answer was unrealistic, the submission was rejected. Supplementary Table S1 presents the questionnaire for assessing dietary habits, based on publicly available information on the UK Biobank website (http://biobank.ctsu.ox.ac.uk/crystal/label.cgi?id=100052). Association tests were conducted using linear regression models for each of the dietary habits, controlling for age, age$^2$, sex, age by sex interaction, age$^2$ by sex interaction, and top 20 principal components. Bread intake and tea intake used in the GWAS have been inverse rank normalized. More detailed information can be found on the website of Neale's lab.

*Major depressive disorder*

The Psychiatric Genomics Consortium (PGC) provided summary statistics from a genome-wide association meta-analysis of MDD, including the core PGC29 cohorts, deCODE cohort, Generation Scotland (GenScot) cohort, Genetic Epidemiology Research on Adult Health and Aging (GERA) cohort, and Integrative Psychiatric Research (iPSYCH) cohort [1]. These cohorts collected data in various places around the world, including the United States, Germany, Australia, Switzerland, Scotland, Europe, the Netherlands, Britain, Ireland, Denmark, Sweden, and Iceland. Cases were identified by structured diagnostic interviews or clinical diagnoses from healthcare electronic records. In total, this meta-analysis included 45,396 MDD patients and 97,250 controls of European ancestry, with 9.8 million genetic variants. More detailed information can be found in Wray et al [1] as well as on the PGC's website (https://www.med.unc.edu/pgc/download-results/).

### Supplementary Tables

#### Supplementary Table S1.

Summary of 20 dietary habits questionnaire

| Trait | Question | Help button information | Notes |
|---|---|---|---|
| Cooked vegetable intake | On average how many heaped tablespoons of COOKED vegetables would you eat per DAY? (Do not include potatoes; put '0' if you do not eat any) | Please provide an average considering your intake over the last year.<br>If you are unsure, please provide an estimate or select Do not know.<br>If you have less than one tablespoon a day select Less than one. | If answer > 50 then rejected<br>-10 represents "Less than one"<br>-1 represents "Do not know"<br>-3 represents "Prefer not to answer" |
| Salad / raw vegetable intake | On average how many heaped tablespoons of SALAD or RAW vegetables would you eat per DAY? (Include lettuce, tomato in sandwiches; put '0' if you do not eat any) | Please provide an average considering your intake over the last year.<br>If you are unsure, please provide an estimate or select Do not know.<br>If you have less than one tablespoon a day select Less than one. | If answer > 50 then rejected<br>-10 represents "Less than one"<br>-1 represents "Do not know"<br>-3 represents "Prefer not to answer" |
| Fresh fruit intake | About how many pieces of FRESH fruit would you eat per DAY? (Count one apple, one banana, 10 grapes etc as one piece; put '0' if you do not eat any) | Please provide an average considering your intake over the last year.<br>If you are unsure, please provide an estimate or select Do not know | If answer > 50 then rejected<br>-10 represents "Less than one"<br>-1 represents "Do not know"<br>-3 represents "Prefer not to answer" |

| Trait | Question | Help button information | Notes |
|---|---|---|---|
| Dried fruit intake | About how many pieces of DRIED fruit would you eat per DAY? (Count one prune, one dried apricot, 10 raisins as one piece; put '0' if you do not eat any) | Please provide an average considering your intake over the last year.<br>If you are unsure, please provide an estimate or select Do not know. | If answer > 100 then rejected<br>-10 represents "Less than one"<br>-1 represents "Do not know"<br>-3 represents "Prefer not to answer" |
| Oily fish intake | How often do you eat oily fish? (e.g. sardines, salmon, mackerel, herring) | Please provide an average considering your intake over the last year.<br>If you are unsure, please provide an estimate or select Do not know.<br>Oily fish include: Salmon Anchovies, Trout Swordfish, Mackerel Bloater, Herring Cacha, Sardines Carp, Pilchards Hilsa, Kipper Jack fish, Eel Katla, Whitebait Orange roughy, Tuna (fresh only) Pangas, Sprats | Options: never, less than once a week, once a week, 2-4 times a week, 5-6 times a week, once or more daily, do not know, prefer not to answer |
| Non-oily fish intake | How often do you eat other types of fish? (e.g. cod, tinned tuna, haddock) | Please provide an average considering your intake over the last year.<br>If you are unsure, please provide an estimate or select Do not know. | Options: never, less than once a week, once a week, 2-4 times a week, 5-6 times a week, once or more daily, do not know, prefer not to answer |

| Trait | Question | Help button information | Notes |
| --- | --- | --- | --- |
| Processed meat intake | How often do you eat processed meats (such as bacon, ham, sausages, meat pies, kebabs, burgers, chicken nuggets)? | Please provide an average considering your intake over the last year<br>If you are unsure, please provide an estimate or select Do not know. | Options: never, less than once a week, once a week, 2-4 times a week, 5-6 times a week, once or more daily, do not know, prefer not to answer |
| Poultry intake | How often do you eat chicken, turkey or other poultry? (Do not count processed meats) | Please provide an average considering your intake over the last year<br>If you are unsure, please provide an estimate or select Do not know. | Options: never, less than once a week, once a week, 2-4 times a week, 5-6 times a week, once or more daily, do not know, prefer not to answer |
| Beef intake | How often do you eat beef? (Do not count processed meats) | Please provide an average considering your intake over the last year<br>If you are unsure, please provide an estimate or select Do not know. | Options: never, less than once a week, once a week, 2-4 times a week, 5-6 times a week, once or more daily, do not know, prefer not to answer |
| Lamb/mutton intake | How often do you eat lamb/mutton? (Do not count processed meats) | Please provide an average considering your intake over the last year<br>If you are unsure, please provide an estimate or select Do not know. | Options: never, less than once a week, once a week, 2-4 times a week, 5-6 times a week, once or more daily, do not know, prefer not to answer |

| Trait | Question | Help button information | Notes |
|---|---|---|---|
| Pork intake | How often do you eat pork? (Do not count processed meats such as bacon or ham) | Please provide an average considering your intake over the last year<br>If you are unsure, please provide an estimate or select Do not know. | Options: never, less than once a week, once a week, 2-4 times a week, 5-6 times a week, once or more daily, do not know, prefer not to answer |
| Cheese intake | How often do you eat cheese? (Include cheese in pizzas, quiches, cheese sauce etc) | Please provide an average considering your intake over the last year<br>If you are unsure, please provide an estimate or select Do not know. | Options: never, less than once a week, once a week, 2-4 times a week, 5-6 times a week, once or more daily, do not know, prefer not to answer |
| Bread intake | How many slices of bread do you eat each WEEK? | For other types of bread:<br>- one bread roll = 2 slices<br>- one pitta bread = 2 slices | If answer < 0 then rejected<br>If answer > 250 then rejected<br>If answer > 50 then participant asked to confirm<br>-10 represents "Less than one"<br>-1 represents "Do not know"<br>-3 represents "Prefer not to answer" |

| Trait | Question | Help button information | Notes |
|---|---|---|---|
| Cereal intake | How many bowls of cereal do you eat a WEEK? | Please provide an average considering your intake over the last year.<br>If you are unsure, please provide an estimate or select Do not know. | If answer < 0 then rejected<br>If answer > 99 then rejected<br>If answer > 14 then participant asked to confirm<br>-10 represents "Less than one"<br>-1 represents "Do not know"<br>-3 represents "Prefer not to answer" |
| Salt added to food | Do you add salt to your food? (Do not include salt used in cooking) | Please provide an average considering your intake over the last year<br>If you are unsure, please provide an estimate or select Do not know. | Options: never/ rarely, sometimes, usually, always, prefer not to answer |
| Tea intake | How many cups of tea do you drink each DAY? (Include black and green tea) | Please provide an average considering your intake over the last year.<br>If you are unsure, please provide an estimate or select Do not know. | If answer < 0 then rejected<br>If answer > 99 then rejected<br>If answer > 20 then participant asked to confirm<br>-10 represents "Less than one"<br>-1 represents "Do not know"<br>-3 represents "Prefer not to answer" |

| Trait | Question | Help button information | Notes |
|---|---|---|---|
| Coffee intake | How many cups of coffee do you drink each DAY? (Include decaffeinated coffee) | Please provide an average considering your intake over the last year.<br>If you are unsure, please provide an estimate or select Do not know. | If answer < 0 then rejected<br>If answer > 99 then rejected<br>If answer > 10 then participant asked to confirm<br>-10 represents "Less than one"<br>-1 represents "Do not know"<br>-3 represents "Prefer not to answer" |
| Hot drink temperature | How do you like your hot drinks? (Such as coffee or tea) | Not available | Options: very hot, hot, warm, do not drink hot drinks, prefer not to answer |
| Water intake | How many glasses of water do you drink each DAY? | Please provide an average considering your intake over the last year.<br>If you are unsure, please provide an estimate or select Do not know. | If answer < 0 then rejected<br>If answer > 99 then rejected<br>If answer > 10 then participant asked to confirm<br>-10 represents "Less than one"<br>-1 represents "Do not know"<br>-3 represents "Prefer not to answer" |
| Alcohol intake frequency | About how often do you drink alcohol? | If this varies a lot, please provide an average considering your intake over the last year | Options: daily or almost daily, three or four times a week, once or twice a week, one to three times a month, special occasions only, never, prefer not to answer |

Note: The information of the questionnaire was extracted from the UK biobank website.

**Supplementary Table S2**

Genome-wide significant SNPs for cooked vegetable intake

| N | SNP | Proxy SNP | $r^2$ for proxy | Effect allele (reference) | β (SE) for cooked vegetable intake | β (SE) for MDD |
|---|---|---|---|---|---|---|
| 1 | rs10156602 | - | - | G(A) | 0.010 (0.002) | -0.027 (0.009) |
| 2 | rs12773105 | - | - | T(C) | 0.010 (0.002) | -0.006 (0.009) |
| 3 | rs204887 | - | - | A(G) | 0.010 (0.002) | 0.003 (0.010) |
| 4 | rs28711392 | - | - | C(T) | -0.010 (0.002) | 0.002 (0.009) |
| 5 | rs6707445 | - | - | A(G) | 0.012 (0.002) | 0.009 (0.009) |
| 6 | rs7619139 | - | - | T(A) | -0.012 (0.002) | -0.020 (0.009) |

SNP, single nucleotide polymorphism; SE, standard error; MDD, major depressive disorder

#### Supplementary Table S3

Genome-wide significant SNPs for salad / raw vegetable intake

| N | SNP | Proxy SNP | $r^2$ for proxy | Effect allele (reference) | β (SE) for salad / raw vegetable intake | β (SE) for MDD |
|---|---|---|---|---|---|---|
| 1 | rs191602006 | - | - | C(T) | 0.018 (0.003) | 0.015 (0.017) |
| 2 | rs200739311 | rs112523595 | 0.85 | T(C) | 0.010 (0.002) | -0.006 (0.010) |
| 3 | rs2517873 | rs9259927 | 0.83 | A(G) | -0.013 (0.002) | -0.019 (0.013) |
| 4 | rs57221424 | - | - | G(C) | 0.011 (0.002) | 0.007 (0.010) |
| 5 | rs572771346 | rs16940676 | 0.82 | T(C) | -0.012 (0.002) | 0.005 (0.011) |
| 6 | rs62461210 | - | - | C(T) | -0.012 (0.002) | -0.014 (0.012) |
| 7 | rs7619139 | - | - | T(A) | -0.013 (0.002) | -0.020 (0.009) |
| 8 | rs790564 | - | - | A(C) | -0.013 (0.002) | -0.022 (0.010) |
| 9 | rs7970482 | - | - | A(G) | -0.010 (0.002) | 0.017 (0.010) |
| 10 | rs946711 | - | - | C(A) | -0.012 (0.002) | 0.002 (0.010) |
| 11 | rs9851987 | - | - | C(T) | 0.010 (0.002) | -0.007 (0.009) |

SNP, single nucleotide polymorphism; SE, standard error; MDD, major depressive disorder

#### Supplementary Table S4

Genome-wide significant SNPs for fresh fruit intake

| N | SNP | Proxy SNP | $r^2$ for proxy | Effect allele (reference) | β (SE) for fresh fruit intake | β (SE) for MDD |
|---|---|---|---|---|---|---|
| 1 | rs10064431 | - | - | T(C) | 0.008 (0.001) | <0.001 (0.009) |
| 2 | rs10249294 | - | - | A(G) | 0.020 (0.001) | 0.010 (0.009) |
| 3 | rs10769936 | - | - | T(C) | 0.009 (0.002) | -0.014 (0.010) |
| 4 | rs11039265 | - | - | A(C) | 0.009 (0.001) | -0.012 (0.010) |
| 5 | rs11074372 | - | - | G(A) | 0.008 (0.001) | -0.001 (0.009) |
| 6 | rs13429231 | - | - | A(G) | 0.008 (0.001) | 0.010 (0.009) |
| 7 | rs145149494 | N/A | N/A | - | - | - |
| 8 | rs1620977 | - | - | A(G) | 0.014 (0.002) | 0.029 (0.011) |
| 9 | rs1698114 | N/A | N/A | - | - | - |
| 10 | rs17258783 | - | - | T(C) | -0.010 (0.002) | <0.001 (0.011) |
| 11 | rs1964272 | - | - | A(G) | 0.009 (0.001) | -0.005 (0.009) |
| 12 | rs2143081 | - | - | G(A) | -0.009 (0.001) | -0.013 (0.009) |
| 13 | rs2246873 | - | - | G(A) | -0.008 (0.001) | <0.001 (0.009) |
| 14 | rs2504671 | - | - | A(C) | 0.013 (0.002) | -0.005 (0.010) |
| 15 | rs2790688 | - | - | T(C) | 0.011 (0.002) | -0.036 (0.013) |
| 16 | rs34162196 | - | - | T(C) | -0.018 (0.002) | -0.028 (0.015) |
| 17 | rs4141843 | - | - | A(T) | 0.009 (0.001) | -0.039 (0.010) |
| 18 | rs429358 | - | - | C(T) | 0.011 (0.002) | -0.040 (0.015) |
| 19 | rs4953149 | - | - | C(T) | -0.009 (0.001) | -0.004 (0.010) |
| 20 | rs5749227 | - | - | A(G) | 0.009 (0.002) | 0.002 (0.011) |
| 21 | rs62298288 | - | - | T(C) | 0.008 (0.001) | -0.016 (0.009) |
| 22 | rs6591836 | - | - | T(C) | 0.008 (0.001) | -0.010 (0.010) |
| 23 | rs66906321 | - | - | T(C) | -0.011 (0.002) | -0.029 (0.012) |
| 24 | rs7072776 | - | - | A(G) | -0.013 (0.002) | 0.003 (0.010) |
| 25 | rs72615728 | - | - | C(G) | 0.008 (0.001) | -0.007 (0.010) |
| 26 | rs73455661 | - | - | G(A) | 0.011 (0.002) | 0.002 (0.010) |
| 27 | rs7916586 | - | - | A(G) | -0.008 (0.001) | 0.012 (0.009) |
| 28 | rs7982441 | - | - | T(C) | 0.009 (0.002) | 0.010 (0.010) |
| 29 | rs8132845 | - | - | T(A) | -0.013 (0.002) | -0.008 (0.019) |
| 30 | rs817223 | - | - | C(T) | -0.008 (0.001) | 0.012 (0.009) |
| 31 | rs862227 | - | - | G(A) | -0.010 (0.001) | -0.020 (0.009) |

SNP, single nucleotide polymorphism; SE, standard error; MDD, major depressive disorder

Notes: rs13069655 and rs2048522 removed for being palindromic with intermediate allele frequencies. N/A indicates that proxy SNP with $r^2 > 0.8$ cannot be found.

#### Supplementary Table S5

Genome-wide significant SNPs for dried fruit intake

| N | SNP | Proxy SNP | $r^2$ for proxy | Effect allele (reference) | β (SE) for dried fruit intake | β (SE) for MDD |
|---|---|---|---|---|---|---|
| 1 | rs10740991 | - | - | G(C) | -0.017 (0.002) | <0.001 (0.010) |
| 2 | rs10765775 | - | - | A(G) | 0.012 (0.002) | -0.011 (0.009) |
| 3 | rs10789340 | - | - | A(G) | -0.015 (0.002) | -0.049 (0.009) |
| 4 | rs10896126 | - | - | G(A) | -0.016 (0.002) | 0.005 (0.010) |
| 5 | rs11586016 | - | - | C(G) | 0.011 (0.002) | 0.001 (0.009) |
| 6 | rs11676282 | - | - | G(A) | 0.011 (0.002) | -0.016 (0.009) |
| 7 | rs11787024 | rs3923107 | 0.99 | C(T) | 0.012 (0.002) | -0.003 (0.010) |
| 8 | rs11899261 | rs13018443 | 0.93 | G(A) | -0.011 (0.002) | 0.003 (0.009) |
| 9 | rs17408272 | - | - | A(G) | -0.012 (0.002) | 0.031 (0.010) |
| 10 | rs1877723 | - | - | T(C) | 0.012 (0.002) | -0.013 (0.010) |
| 11 | rs2328887 | - | - | T(C) | -0.019 (0.003) | -0.025 (0.015) |
| 12 | rs2422013 | - | - | T(G) | 0.011 (0.002) | -0.021 (0.009) |
| 13 | rs2510344 | - | - | C(T) | 0.013 (0.002) | 0.022 (0.009) |
| 14 | rs2857597 | N/A | N/A | - | - | - |
| 15 | rs34162196 | - | - | T(C) | -0.023 (0.003) | -0.028 (0.015) |
| 16 | rs35164474 | - | - | G(T) | 0.012 (0.002) | 0.003 (0.010) |
| 17 | rs429358 | - | - | C(T) | 0.023 (0.003) | -0.040 (0.015) |
| 18 | rs4532156 | - | - | T(C) | 0.013 (0.002) | -0.021 (0.010) |
| 19 | rs4868800 | - | - | G(T) | -0.011 (0.002) | 0.027 (0.009) |
| 20 | rs55950910 | - | - | T(A) | -0.020 (0.003) | 0.017 (0.015) |
| 21 | rs61937394 | - | - | G(T) | -0.015 (0.002) | -0.001 (0.012) |
| 22 | rs62444881 | - | - | T(C) | 0.019 (0.002) | -0.038 (0.011) |
| 23 | rs6545770 | - | - | A(T) | 0.012 (0.002) | -0.031 (0.011) |
| 24 | rs7599488 | - | - | T(C) | -0.011 (0.002) | -0.002 (0.009) |
| 25 | rs898751 | - | - | T(C) | -0.011 (0.002) | -0.004 (0.009) |
| 26 | rs924762 | - | - | T(C) | 0.012 (0.002) | -0.015 (0.010) |
| 27 | rs9385269 | - | - | C(T) | -0.012 (0.002) | -0.003 (0.009) |
| 28 | rs9729959 | - | - | T(C) | 0.013 (0.002) | -0.002 (0.011) |

SNP, single nucleotide polymorphism; SE, standard error; MDD, major depressive disorder

Notes: N/A indicates that proxy SNP with $r^2 > 0.8$ cannot be found.

**Supplementary Table S6**

Genome-wide significant SNPs for oily fish intake

| N | SNP | Proxy SNP | $r^2$ for proxy | Effect allele (reference) | β (SE) for oily fish intake | β (SE) for MDD |
|---|---|---|---|---|---|---|
| 1 | rs10267710 | - | - | T(G) | -0.014 (0.002) | 0.010 (0.009) |
| 2 | rs11233632 | - | - | C(T) | 0.013 (0.002) | -0.001 (0.009) |
| 3 | rs114227716 | - | - | G(A) | 0.050 (0.009) | 0.082 (0.051) |
| 4 | rs114497213 | - | - | T(G) | 0.027 (0.005) | 0.004 (0.022) |
| 5 | rs11692742 | - | - | A(G) | -0.013 (0.002) | 0.013 (0.011) |
| 6 | rs11767283 | - | - | G(A) | 0.019 (0.003) | 0.003 (0.012) |
| 7 | rs11859365 | - | - | C(A) | 0.024 (0.002) | -0.006 (0.011) |
| 8 | rs1217101 | - | - | C(G) | -0.014 (0.003) | -0.028 (0.011) |
| 9 | rs12576625 | - | - | T(C) | -0.017 (0.003) | 0.008 (0.012) |
| 10 | rs12787863 | - | - | T(C) | -0.013 (0.002) | 0.016 (0.009) |
| 11 | rs1371272 | - | - | A(G) | 0.017 (0.003) | 0.015 (0.013) |
| 12 | rs1449404 | - | - | G(C) | 0.016 (0.003) | -0.003 (0.011) |
| 13 | rs1876245 | - | - | C(T) | 0.013 (0.002) | -0.006 (0.009) |
| 14 | rs191728525 | - | - | G(T) | 0.015 (0.002) | -0.013 (0.010) |
| 15 | rs2026092 | - | - | A(G) | -0.022 (0.002) | 0.002 (0.010) |
| 16 | rs2287922 | - | - | G(A) | 0.018 (0.002) | -0.005 (0.009) |
| 17 | rs2315507 | - | - | G(A) | -0.014 (0.002) | 0.009 (0.009) |
| 18 | rs2413045 | - | - | A(G) | 0.020 (0.003) | 0.007 (0.012) |
| 19 | rs242641 | - | - | T(C) | -0.013 (0.002) | -0.006 (0.010) |
| 20 | rs28533540 | - | - | G(A) | -0.016 (0.002) | 0.014 (0.009) |
| 21 | rs3124402 | - | - | A(G) | 0.020 (0.002) | 0.009 (0.010) |
| 22 | rs3132934 | N/A | N/A | - | - | - |
| 23 | rs3136469 | - | - | A(T) | 0.022 (0.004) | 0.015 (0.016) |
| 24 | rs35287743 | - | - | T(G) | -0.026 (0.003) | -0.016 (0.014) |
| 25 | rs3734543 | - | - | C(G) | -0.024 (0.003) | -0.063 (0.015) |
| 26 | rs45501495 | - | - | T(C) | 0.018 (0.003) | <0.001 (0.011) |
| 27 | rs4869859 | - | - | C(T) | 0.016 (0.002) | 0.002 (0.009) |
| 28 | rs552234 | - | - | A(G) | -0.013 (0.002) | 0.013 (0.009) |
| 29 | rs56094641 | - | - | G(A) | 0.018 (0.002) | 0.016 (0.009) |
| 30 | rs6089752 | N/A | N/A | - | - | - |
| 31 | rs62148582 | - | - | G(A) | -0.012 (0.002) | -0.003 (0.009) |
| 32 | rs6667502 | - | - | C(T) | 0.012 (0.002) | <0.001 (0.009) |
| 33 | rs7225002 | N/A | N/A | - | - | - |
| 34 | rs75887709 | - | - | G(A) | -0.017 (0.003) | -0.025 (0.014) |

| N | SNP | Proxy SNP | $r^2$ for proxy | Effect allele (reference) | β (SE) for oily fish intake | β (SE) for MDD |
|---|---|---|---|---|---|---|
| 35 | rs875257 | - | - | A(G) | 0.013 (0.002) | 0.004 (0.010) |
| 36 | rs9841174 | - | - | C(T) | 0.015 (0.002) | -0.009 (0.009) |

SNP, single nucleotide polymorphism; SE, standard error; MDD, major depressive disorder

Notes: rs11986122 and rs67474621 removed for being palindromic with intermediate allele frequencies. N/A indicates that proxy SNP with $r^2 > 0.8$ cannot be found.

#### Supplementary Table S7

Genome-wide significant SNPs for non-oily fish intake

| N | SNP | Proxy SNP | $r^2$ for proxy | Effect allele (reference) | β (SE) for non-oily fish intake | β (SE) for MDD |
|---|---|---|---|---|---|---|
| 1 | rs2447091 | - | - | C(T) | -0.011 (0.002) | -0.003 (0.010) |
| 2 | rs3129981 | N/A | N/A | - | - | - |
| 3 | rs35287743 | - | - | T(G) | -0.016 (0.003) | -0.016 (0.014) |
| 4 | rs35493868 | - | - | G(C) | -0.013 (0.002) | -0.015 (0.012) |
| 5 | rs56094641 | - | - | G(A) | 0.012 (0.002) | 0.016 (0.009) |
| 6 | rs62408273 | - | - | T(C) | -0.024 (0.004) | -0.020 (0.017) |
| 7 | rs8103840 | - | - | T(C) | 0.016 (0.002) | <0.001 (0.009) |

SNP, single nucleotide polymorphism; SE, standard error; MDD, major depressive disorder

Note: N/A indicates that proxy SNP with $r^2 > 0.8$ cannot be found.

#### Supplementary Table S8

Genome-wide significant SNPs for processed meat intake

| N | SNP | Proxy SNP | $r^2$ for proxy | Effect allele (reference) | β (SE) for processed meat intake | β (SE) for MDD |
|---|---|---|---|---|---|---|
| 1 | rs10454812 | - | - | C(A) | -0.022 (0.004) | -0.020 (0.016) |
| 2 | rs12127789 | - | - | T(G) | -0.022 (0.004) | 0.020 (0.014) |
| 3 | rs13091492 | - | - | G(A) | 0.014 (0.002) | -0.006 (0.009) |
| 4 | rs137831 | - | - | C(A) | -0.018 (0.003) | -0.004 (0.011) |
| 5 | rs158582 | - | - | T(C) | -0.016 (0.003) | 0.011 (0.010) |
| 6 | rs35399883 | - | - | C(G) | 0.014 (0.003) | -0.023 (0.010) |
| 7 | rs838133 | - | - | A(G) | -0.019 (0.002) | -0.002 (0.010) |
| 8 | rs9809856 | - | - | G(A) | 0.014 (0.002) | 0.009 (0.009) |

SNP, single nucleotide polymorphism; SE, standard error; MDD, major depressive disorder

Note: rs7843470 removed for being palindromic with intermediate allele frequencies.

**Supplementary Table S9**

Genome-wide significant SNPs for poultry intake

| N | SNP | Proxy SNP | $r^2$ for proxy | Effect allele (reference) | β (SE) for poultry intake | β (SE) for MDD |
|---|---|---|---|---|---|---|
| 1 | rs1062633 | - | - | T(C) | -0.011 (0.002) | -0.020 (0.009) |
| 2 | rs2975736 | - | - | T(G) | 0.017 (0.003) | 0.014 (0.013) |
| 3 | rs7829800 | - | - | A(G) | -0.012 (0.002) | -0.004 (0.010) |

SNP, single nucleotide polymorphism; SE, standard error; MDD, major depressive disorder

#### Supplementary Table S10

Genome-wide significant SNPs for beef intake

| N | SNP | Proxy SNP | $r^2$ for proxy | Effect allele (reference) | β (SE) for beef intake | β (SE) for MDD |
|---|---|---|---|---|---|---|
| 1 | rs10739743 | - | - | C(A) | -0.012 (0.002) | 0.002 (0.010) |
| 2 | rs10962119 | - | - | T(C) | -0.013 (0.002) | -0.022 (0.010) |
| 3 | rs11720703 | - | - | T(C) | 0.014 (0.002) | -0.016 (0.009) |
| 4 | rs12483998 | - | - | G(A) | -0.015 (0.002) | -0.005 (0.012) |
| 5 | rs17817288 | - | - | G(A) | -0.012 (0.002) | 0.018 (0.009) |
| 6 | rs2815753 | - | - | G(A) | 0.015 (0.002) | -0.048 (0.009) |
| 7 | rs61853274 | - | - | A(G) | -0.014 (0.002) | -0.043 (0.013) |
| 8 | rs62396185 | - | - | C(G) | -0.016 (0.002) | 0.024 (0.010) |
| 9 | rs7240986 | - | - | A(G) | -0.011 (0.002) | 0.027 (0.009) |

SNP, single nucleotide polymorphism; SE, standard error; MDD, major depressive disorder

**Supplementary Table S11**

Genome-wide significant SNPs for lamb/mutton intake

| N | SNP | Proxy SNP | $r^2$ for proxy | Effect allele (reference) | β (SE) for lamb/mutton intake | β (SE) for MDD |
|---|---|---|---|---|---|---|
| 1 | rs11090045 | - | - | A(G) | -0.010 (0.002) | 0.008 (0.010) |
| 2 | rs13069895 | rs67928440 | 1.00 | C(A) | -0.011 (0.002) | 0.019 (0.010) |
| 3 | rs1698114 | N/A | N/A | - | - | - |
| 4 | rs2222760 | - | - | A(G) | -0.010 (0.002) | -0.006 (0.010) |
| 5 | rs2678900 | - | - | G(T) | 0.011 (0.002) | 0.010 (0.009) |
| 6 | rs276447 | - | - | T(C) | -0.013 (0.002) | -0.016 (0.009) |
| 7 | rs2815753 | - | - | G(A) | 0.010 (0.002) | -0.048 (0.009) |
| 8 | rs3909727 | - | - | A(G) | -0.012 (0.002) | 0.008 (0.012) |
| 9 | rs3964074 | - | - | T(C) | 0.009 (0.002) | 0.005 (0.009) |
| 10 | rs429358 | - | - | C(T) | -0.015 (0.002) | -0.040 (0.015) |
| 11 | rs58330009 | N/A | N/A | - | - | - |
| 12 | rs62106258 | - | - | C(T) | 0.022 (0.004) | 0.007 (0.023) |
| 13 | rs677679 | - | - | C(T) | 0.010 (0.002) | 0.022 (0.009) |
| 14 | rs75222322 | - | - | A(C) | -0.013 (0.002) | 0.006 (0.013) |
| 15 | rs8053361 | - | - | T(C) | 0.013 (0.002) | -0.009 (0.011) |
| 16 | rs9527311 | rs1998693 | 1.00 | T(A) | 0.010 (0.002) | 0.015 (0.009) |

SNP, single nucleotide polymorphism; SE, standard error; MDD, major depressive disorder

Note: N/A indicates that proxy SNP with $r^2 > 0.8$ cannot be found.

**Supplementary Table S12**

Genome-wide significant SNPs for pork intake

| N | SNP | Proxy SNP | $r^2$ for proxy | Effect allele (reference) | β (SE) for pork intake | β (SE) for MDD |
|---|---|---|---|---|---|---|
| 1 | rs10972033 | - | - | T(G) | 0.010 (0.002) | -0.001 (0.009) |
| 2 | rs12103229 | - | - | C(A) | 0.010 (0.002) | 0.005 (0.009) |
| 3 | rs1355171 | - | - | A(C) | -0.009 (0.002) | -0.018 (0.009) |
| 4 | rs429358 | - | - | C(T) | -0.014 (0.002) | -0.040 (0.015) |
| 5 | rs6420102 | N/A | N/A | - | - | - |
| 6 | rs9379832 | - | - | G(A) | -0.012 (0.002) | 0.019 (0.011) |

SNP, single nucleotide polymorphism; SE, standard error; MDD, major depressive disorder

Note: N/A indicates that proxy SNP with $r^2 > 0.8$ cannot be found.

**Supplementary Table S13**

Genome-wide significant SNPs for cheese intake

| N | SNP | Proxy SNP | $r^2$ for proxy | Effect allele (reference) | β (SE) for cheese intake | β (SE) for MDD |
|---|---|---|---|---|---|---|
| 1 | rs1073242 | - | - | G(A) | -0.014 (0.003) | 0.009 (0.009) |
| 2 | rs10861798 | - | - | A(G) | 0.014 (0.003) | 0.011 (0.010) |
| 3 | rs10938397 | - | - | G(A) | -0.015 (0.003) | 0.011 (0.009) |
| 4 | rs117572343 | N/A | N/A | - | - | - |
| 5 | rs1291145 | - | - | T(C) | 0.021 (0.003) | -0.002 (0.010) |
| 6 | rs13107325 | - | - | T(C) | -0.028 (0.005) | -0.027 (0.019) |
| 7 | rs1322537 | - | - | C(T) | 0.022 (0.003) | -0.011 (0.012) |
| 8 | rs144216645 | N/A | N/A | - | - | - |
| 9 | rs151180 | rs34839 | 0.97 | T(G) | -0.016 (0.003) | -0.001 (0.009) |
| 10 | rs1783826 | - | - | T(G) | 0.016 (0.003) | -0.022 (0.009) |
| 11 | rs1931814 | - | - | A(G) | -0.016 (0.003) | 0.006 (0.009) |
| 12 | rs1984442 | - | - | T(C) | 0.015 (0.003) | 0.016 (0.009) |
| 13 | rs2339928 | - | - | G(A) | -0.016 (0.003) | 0.008 (0.010) |
| 14 | rs2515649 | - | - | A(G) | 0.018 (0.003) | -0.006 (0.010) |
| 15 | rs2960578 | - | - | G(T) | 0.020 (0.003) | 0.022 (0.009) |
| 16 | rs34198643 | - | - | T(C) | -0.018 (0.003) | 0.025 (0.011) |
| 17 | rs504675 | - | - | T(C) | 0.028 (0.003) | -0.005 (0.009) |
| 18 | rs60198071 | - | - | G(A) | 0.017 (0.003) | -0.008 (0.010) |
| 19 | rs62236533 | - | - | A(G) | 0.029 (0.004) | -0.017 (0.016) |
| 20 | rs641811 | - | - | A(G) | 0.017 (0.003) | -0.027 (0.011) |
| 21 | rs6774906 | - | - | C(A) | 0.036 (0.006) | -0.031 (0.025) |
| 22 | rs7012814 | - | - | A(G) | -0.019 (0.003) | -0.002 (0.009) |
| 23 | rs73024305 | - | - | C(G) | 0.033 (0.006) | -0.010 (0.019) |
| 24 | rs7439876 | - | - | G(A) | -0.014 (0.003) | 0.010 (0.009) |
| 25 | rs76632611 | rs12447542 | 0.92 | T(C) | 0.021 (0.004) | 0.019 (0.014) |
| 26 | rs77742462 | - | - | G(A) | -0.051 (0.009) | 0.016 (0.030) |
| 27 | rs78370515 | - | - | T(C) | -0.034 (0.006) | 0.004 (0.021) |
| 28 | rs78876700 | - | - | A(G) | 0.022 (0.004) | -0.013 (0.014) |
| 29 | rs7936836 | - | - | A(C) | 0.015 (0.003) | 0.016 (0.009) |
| 30 | rs919109 | - | - | C(G) | 0.020 (0.004) | -0.019 (0.013) |
| 31 | rs933738 | - | - | G(A) | 0.019 (0.003) | 0.022 (0.012) |
| 32 | rs9649582 | - | - | T(A) | -0.017 (0.003) | 0.014 (0.010) |

SNP, single nucleotide polymorphism; SE, standard error; MDD, major depressive disorder

Note: rs4579080 removed for being palindromic with intermediate allele frequencies. N/A indicates that proxy SNP with $r^2 > 0.8$ cannot be found.

**Supplementary Table S14**

Genome-wide significant SNPs for bread intake

| N | SNP | Proxy SNP | $r^2$ for proxy | Effect allele (reference) | β (SE) for bread intake | β (SE) for MDD |
|---|---|---|---|---|---|---|
| 1 | rs11060853 | - | - | G(A) | -0.014 (0.002) | -0.004 (0.010) |
| 2 | rs115877304 | - | - | T(C) | 0.031 (0.005) | -0.034 (0.024) |
| 3 | rs11620650 | - | - | C(T) | -0.014 (0.002) | 0.004 (0.009) |
| 4 | rs11691161 | - | - | G(A) | -0.013 (0.002) | 0.006 (0.009) |
| 5 | rs13016665 | - | - | A(C) | 0.013 (0.002) | 0.019 (0.009) |
| 6 | rs182549 | - | - | C(T) | 0.017 (0.003) | -0.015 (0.011) |
| 7 | rs2068650 | - | - | C(A) | -0.013 (0.002) | 0.019 (0.009) |
| 8 | rs4665972 | - | - | T(C) | 0.016 (0.002) | -0.003 (0.009) |
| 9 | rs62394558 | - | - | A(G) | -0.013 (0.002) | 0.027 (0.009) |
| 10 | rs6464566 | - | - | A(G) | -0.024 (0.002) | 0.011 (0.009) |
| 11 | rs656817 | - | - | G(A) | -0.014 (0.002) | -0.022 (0.010) |
| 12 | rs7134238 | - | - | G(T) | 0.014 (0.002) | -0.013 (0.009) |
| 13 | rs7647649 | - | - | T(C) | 0.019 (0.002) | 0.012 (0.010) |
| 14 | rs7965658 | - | - | A(G) | 0.021 (0.003) | 0.020 (0.011) |
| 15 | rs9440302 | - | - | A(G) | -0.014 (0.002) | -0.019 (0.009) |
| 16 | rs9877994 | - | - | G(A) | -0.015 (0.002) | -0.002 (0.009) |

SNP, single nucleotide polymorphism; SE, standard error; MDD, major depressive disorder

#### Supplementary Table S15

Genome-wide significant SNPs for cereal intake

| N | SNP | Proxy SNP | $r^2$ for proxy | Effect allele (reference) | β (SE) for cereal intake | β (SE) for MDD |
|---|---|---|---|---|---|---|
| 1 | rs11940694 | - | - | A(G) | 0.012 (0.002) | -0.005 (0.009) |
| 2 | rs12713415 | - | - | G(C) | 0.015 (0.002) | -0.024 (0.010) |
| 3 | rs1609783 | - | - | G(A) | 0.016 (0.002) | -0.018 (0.009) |
| 4 | rs2147044 | - | - | A(G) | 0.013 (0.002) | -0.015 (0.011) |
| 5 | rs2278757 | - | - | A(G) | 0.012 (0.002) | -0.011 (0.009) |
| 6 | rs2465018 | - | - | A(G) | 0.019 (0.002) | -0.007 (0.011) |
| 7 | rs2470893 | - | - | T(C) | -0.015 (0.002) | 0.005 (0.010) |
| 8 | rs2725371 | - | - | A(G) | -0.013 (0.002) | 0.006 (0.010) |
| 9 | rs2799849 | - | - | C(T) | 0.014 (0.002) | -0.016 (0.010) |
| 10 | rs2817377 | - | - | G(A) | -0.012 (0.002) | -0.010 (0.009) |
| 11 | rs4953153 | - | - | A(G) | -0.011 (0.002) | 0.006 (0.009) |
| 12 | rs544372 | - | - | T(C) | -0.011 (0.002) | -0.005 (0.009) |
| 13 | rs55826424 | N/A | N/A | - | - | - |
| 14 | rs56131196 | - | - | A(G) | 0.018 (0.003) | -0.029 (0.012) |
| 15 | rs56259105 | - | - | C(T) | 0.015 (0.003) | -0.031 (0.012) |
| 16 | rs68136852 | - | - | A(C) | -0.017 (0.003) | 0.024 (0.013) |
| 17 | rs7113478 | - | - | A(G) | -0.012 (0.002) | 0.013 (0.009) |
| 18 | rs7650602 | - | - | C(T) | 0.012 (0.002) | -0.011 (0.009) |
| 19 | rs8097544 | - | - | G(A) | -0.026 (0.003) | 0.020 (0.013) |
| 20 | rs838133 | - | - | A(G) | 0.020 (0.002) | -0.002 (0.010) |
| 21 | rs9374896 | - | - | T(C) | 0.017 (0.002) | -0.001 (0.009) |
| 22 | rs993354 | - | - | A(T) | 0.013 (0.002) | -0.007 (0.009) |

SNP, single nucleotide polymorphism; SE, standard error; MDD, major depressive disorder

Notes: rs1965342 removed for being palindromic with intermediate allele frequencies. N/A indicates that proxy SNP with $r^2 > 0.8$ cannot be found.

**Supplementary Table S16**

Genome-wide significant SNPs for salt added to food

| N | SNP | Proxy SNP | $r^2$ for proxy | Effect allele (reference) | β (SE) for salt added to food | β (SE) for MDD |
|---|---|---|---|---|---|---|
| 1 | rs1008078 | - | - | T(C) | 0.012 (0.002) | 0.011 (0.009) |
| 2 | rs1045411 | - | - | T(C) | -0.015 (0.002) | 0.010 (0.010) |
| 3 | rs10736951 | - | - | G(C) | 0.012 (0.002) | 0.002 (0.009) |
| 4 | rs10752999 | - | - | A(C) | -0.012 (0.002) | -0.010 (0.010) |
| 5 | rs10971930 | - | - | C(T) | 0.018 (0.003) | -0.006 (0.013) |
| 6 | rs10994732 | - | - | T(C) | 0.018 (0.002) | 0.008 (0.009) |
| 7 | rs11126666 | - | - | A(G) | -0.013 (0.002) | 0.003 (0.010) |
| 8 | rs11210985 | - | - | A(G) | -0.016 (0.002) | 0.007 (0.009) |
| 9 | rs113271874 | rs147896014 | 1.00 | T(C) | -0.013 (0.002) | 0.002 (0.010) |
| 10 | rs12499144 | - | - | C(A) | -0.014 (0.002) | 0.005 (0.009) |
| 11 | rs12501838 | - | - | T(A) | 0.014 (0.002) | -0.010 (0.011) |
| 12 | rs12579997 | - | - | C(G) | 0.016 (0.003) | -0.002 (0.012) |
| 13 | rs12658060 | - | - | C(T) | -0.026 (0.002) | -0.004 (0.011) |
| 14 | rs12947874 | - | - | G(A) | 0.013 (0.002) | 0.013 (0.011) |
| 15 | rs13028667 | - | - | A(G) | 0.017 (0.003) | 0.002 (0.013) |
| 16 | rs13131880 | - | - | C(T) | -0.014 (0.002) | 0.010 (0.011) |
| 17 | rs1339157 | - | - | C(T) | -0.014 (0.002) | -0.004 (0.009) |
| 18 | rs13420169 | - | - | C(A) | 0.021 (0.002) | 0.017 (0.011) |
| 19 | rs1409294 | - | - | T(C) | 0.012 (0.002) | 0.008 (0.009) |
| 20 | rs147831713 | rs76025409 | 0.98 | C(A) | 0.014 (0.002) | 0.057 (0.010) |
| 21 | rs1521193 | - | - | A(G) | 0.012 (0.002) | 0.017 (0.009) |
| 22 | rs1538172 | - | - | G(A) | -0.011 (0.002) | -0.025 (0.009) |
| 23 | rs1584336 | - | - | A(G) | -0.027 (0.002) | -0.005 (0.012) |
| 24 | rs1726866 | - | - | G(A) | -0.027 (0.002) | -0.001 (0.009) |
| 25 | rs17805497 | - | - | C(T) | 0.014 (0.002) | -0.009 (0.010) |
| 26 | rs2053682 | - | - | C(A) | 0.017 (0.002) | 0.002 (0.010) |
| 27 | rs2165127 | - | - | A(G) | 0.015 (0.003) | 0.028 (0.012) |
| 28 | rs2263636 | - | - | A(C) | -0.014 (0.002) | -0.003 (0.010) |
| 29 | rs2466184 | - | - | A(T) | 0.017 (0.003) | 0.013 (0.013) |
| 30 | rs2687169 | - | - | T(C) | 0.015 (0.002) | <0.001 (0.009) |
| 31 | rs2852348 | - | - | G(A) | -0.013 (0.002) | 0.004 (0.009) |
| 32 | rs34906832 | - | - | G(A) | 0.016 (0.003) | 0.011 (0.013) |
| 33 | rs35099536 | - | - | C(A) | 0.027 (0.004) | 0.019 (0.017) |
| 34 | rs400750 | - | - | T(G) | -0.017 (0.002) | -0.007 (0.009) |

| N | SNP | Proxy SNP | $r^2$ for proxy | Effect allele (reference) | β (SE) for salt added to food | β (SE) for MDD |
|---|---|---|---|---|---|---|
| 35 | rs413172 | rs273617 | 0.91 | C(T) | -0.014 (0.002) | 0.011 (0.010) |
| 36 | rs429358 | - | - | C(T) | -0.016 (0.003) | -0.040 (0.015) |
| 37 | rs4580876 | - | - | A(G) | -0.012 (0.002) | -0.004 (0.009) |
| 38 | rs4739105 | - | - | T(C) | -0.014 (0.002) | -0.028 (0.011) |
| 39 | rs4856591 | - | - | T(G) | 0.018 (0.002) | -0.008 (0.010) |
| 40 | rs510901 | - | - | T(C) | 0.014 (0.002) | 0.003 (0.009) |
| 41 | rs528301 | - | - | G(A) | -0.016 (0.002) | 0.006 (0.009) |
| 42 | rs55897719 | - | - | A(C) | 0.013 (0.002) | 0.015 (0.010) |
| 43 | rs62098445 | - | - | A(C) | -0.018 (0.002) | -0.013 (0.010) |
| 44 | rs6443950 | - | - | A(T) | 0.015 (0.002) | -0.024 (0.009) |
| 45 | rs6707445 | - | - | A(G) | 0.012 (0.002) | 0.009 (0.009) |
| 46 | rs6795756 | - | - | G(A) | 0.015 (0.002) | 0.030 (0.009) |
| 47 | rs6887291 | - | - | T(G) | -0.012 (0.002) | -0.003 (0.009) |
| 48 | rs7232972 | rs7505519 | 0.99 | G(A) | 0.013 (0.002) | 0.021 (0.009) |
| 49 | rs72748335 | - | - | G(A) | -0.012 (0.002) | -0.025 (0.010) |
| 50 | rs7591518 | - | - | C(T) | -0.020 (0.002) | 0.026 (0.010) |
| 51 | rs7670308 | - | - | A(G) | -0.013 (0.002) | -0.019 (0.009) |
| 52 | rs76766085 | - | - | T(C) | 0.026 (0.003) | 0.034 (0.015) |
| 53 | rs7803164 | - | - | T(C) | -0.016 (0.002) | -0.012 (0.010) |
| 54 | rs8022455 | - | - | T(C) | 0.012 (0.002) | <0.001 (0.009) |
| 55 | rs868720 | - | - | C(G) | 0.017 (0.002) | 0.001 (0.010) |
| 56 | rs9404418 | rs4133417 | 0.90 | G(A) | -0.015 (0.002) | 0.006 (0.009) |
| 57 | rs949880 | - | - | G(A) | 0.013 (0.002) | -0.003 (0.009) |
| 58 | rs9539965 | - | - | G(A) | -0.013 (0.002) | 0.010 (0.010) |
| 59 | rs961044 | - | - | C(T) | 0.016 (0.003) | 0.020 (0.013) |
| 60 | rs9611875 | N/A | N/A | - | - | - |
| 61 | rs9852600 | - | - | T(C) | 0.018 (0.003) | -0.013 (0.012) |
| 62 | rs9971382 | - | - | G(T) | -0.015 (0.002) | 0.011 (0.009) |

SNP, single nucleotide polymorphism; SE, standard error; MDD, major depressive disorder

Notes: N/A indicates that proxy SNP with $r^2 > 0.8$ cannot be found.

**Supplementary Table S17**

Genome-wide significant SNPs for tea intake

| N | SNP | Proxy SNP | $r^2$ for proxy | Effect allele (reference) | β (SE) for tea intake | β (SE) for MDD |
|---|---|---|---|---|---|---|
| 1 | rs11022751 | - | - | C(T) | 0.017 (0.003) | -0.013 (0.010) |
| 2 | rs11204711 | rs11204708 | 1.00 | G(A) | 0.015 (0.002) | 0.008 (0.011) |
| 3 | rs11487328 | rs10912824 | 0.97 | C(G) | -0.015 (0.002) | 0.002 (0.009) |
| 4 | rs1156588 | - | - | G(A) | -0.016 (0.003) | -0.006 (0.011) |
| 5 | rs12591786 | - | - | T(C) | -0.021 (0.003) | -0.002 (0.013) |
| 6 | rs1481012 | - | - | G(A) | -0.026 (0.004) | 0.009 (0.014) |
| 7 | rs182050989 | - | - | T(C) | -0.040 (0.007) | 0.007 (0.022) |
| 8 | rs185115295 | rs150329203 | 1.00 | C(T) | -0.018 (0.003) | 0.006 (0.010) |
| 9 | rs2071207 | - | - | C(T) | -0.013 (0.002) | -0.014 (0.009) |
| 10 | rs2117137 | - | - | G(A) | 0.014 (0.002) | 0.015 (0.009) |
| 11 | rs2273447 | - | - | T(A) | 0.020 (0.003) | -0.017 (0.011) |
| 12 | rs2465018 | - | - | A(G) | 0.023 (0.003) | -0.007 (0.011) |
| 13 | rs2472297 | - | - | T(C) | 0.054 (0.003) | 0.011 (0.010) |
| 14 | rs4410790 | - | - | T(C) | -0.039 (0.002) | 0.001 (0.009) |
| 15 | rs4808940 | - | - | G(C) | 0.015 (0.003) | -0.019 (0.009) |
| 16 | rs4817505 | - | - | C(T) | 0.015 (0.002) | -0.010 (0.010) |
| 17 | rs57292194 | - | - | C(T) | 0.026 (0.005) | 0.029 (0.019) |
| 18 | rs6462899 | - | - | T(A) | -0.014 (0.002) | 0.008 (0.009) |
| 19 | rs6467958 | - | - | T(C) | 0.023 (0.003) | -0.016 (0.010) |
| 20 | rs713598 | - | - | G(C) | 0.014 (0.002) | 0.003 (0.009) |
| 21 | rs73424602 | - | - | T(C) | -0.015 (0.002) | 0.006 (0.009) |
| 22 | rs9624470 | - | - | G(A) | -0.025 (0.002) | 0.012 (0.009) |
| 23 | rs977474 | - | - | C(T) | -0.021 (0.003) | 0.009 (0.013) |

SNP, single nucleotide polymorphism; SE, standard error; MDD, major depressive disorder

#### Supplementary Table S18

Genome-wide significant SNPs for coffee intake

| N | SNP | Proxy SNP | $r^2$ for proxy | Effect allele (reference) | β (SE) for coffee intake | β (SE) for MDD |
|---|---|---|---|---|---|---|
| 1 | rs10127720 | - | - | T(C) | -0.014 (0.002) | 0.007 (0.010) |
| 2 | rs10489219 | - | - | A(G) | 0.018 (0.003) | 0.015 (0.015) |
| 3 | rs1057868 | - | - | T(C) | 0.021 (0.002) | -0.017 (0.010) |
| 4 | rs12514566 | - | - | A(G) | -0.011 (0.002) | 0.009 (0.010) |
| 5 | rs1260326 | - | - | T(C) | -0.014 (0.002) | <0.001 (0.009) |
| 6 | rs13271359 | - | - | T(C) | -0.012 (0.002) | -0.011 (0.011) |
| 7 | rs13387090 | - | - | T(G) | -0.017 (0.002) | -0.030 (0.013) |
| 8 | rs2465037 | - | - | A(C) | -0.011 (0.002) | -0.001 (0.009) |
| 9 | rs2472297 | - | - | T(C) | 0.047 (0.002) | 0.011 (0.010) |
| 10 | rs34060476 | - | - | G(A) | 0.020 (0.003) | -0.011 (0.014) |
| 11 | rs3814424 | - | - | T(C) | 0.020 (0.003) | 0.029 (0.012) |
| 12 | rs4410790 | - | - | T(C) | -0.039 (0.002) | 0.001 (0.009) |
| 13 | rs4925114 | - | - | A(G) | -0.012 (0.002) | -0.007 (0.010) |
| 14 | rs56094641 | - | - | G(A) | 0.019 (0.002) | 0.016 (0.009) |
| 15 | rs56113850 | - | - | T(C) | -0.013 (0.002) | 0.019 (0.010) |
| 16 | rs570263 | - | - | T(C) | 0.011 (0.002) | -0.011 (0.010) |
| 17 | rs5760378 | - | - | C(A) | -0.015 (0.002) | 0.005 (0.011) |
| 18 | rs57918684 | - | - | A(G) | 0.014 (0.003) | -0.009 (0.013) |
| 19 | rs597045 | - | - | T(A) | -0.011 (0.002) | -0.010 (0.010) |
| 20 | rs6062682 | - | - | T(C) | 0.011 (0.002) | -0.012 (0.009) |
| 21 | rs66723169 | - | - | A(C) | 0.017 (0.002) | -0.010 (0.011) |
| 22 | rs67595273 | - | - | A(C) | 0.014 (0.002) | 0.014 (0.012) |
| 23 | rs73075167 | - | - | T(A) | -0.016 (0.003) | 0.010 (0.014) |
| 24 | rs7811609 | - | - | T(C) | 0.010 (0.002) | -0.011 (0.009) |
| 25 | rs9398171 | - | - | C(T) | -0.012 (0.002) | -0.021 (0.010) |

SNP, single nucleotide polymorphism; SE, standard error; MDD, major depressive disorder

**Supplementary Table S19**

Genome-wide significant SNPs for hot drink temperature

| N | SNP | Proxy SNP | $r^2$ for proxy | Effect allele (reference) | β (SE) for hot drink temperature | β (SE) for MDD |
|---|---|---|---|---|---|---|
| 1 | rs10273733 | - | - | T(C) | -0.009 (0.001) | -0.024 (0.010) |
| 2 | rs10764990 | - | - | G(A) | -0.010 (0.001) | -0.006 (0.010) |
| 3 | rs10784379 | - | - | C(T) | -0.009 (0.001) | 0.024 (0.009) |
| 4 | rs10829603 | - | - | G(T) | 0.008 (0.001) | -0.018 (0.009) |
| 5 | rs10927006 | - | - | C(T) | -0.014 (0.002) | -0.002 (0.013) |
| 6 | rs1144428 | - | - | G(A) | -0.012 (0.002) | 0.001 (0.011) |
| 7 | rs11710570 | - | - | C(T) | -0.008 (0.001) | -0.011 (0.009) |
| 8 | rs12429545 | - | - | A(G) | -0.013 (0.002) | 0.024 (0.014) |
| 9 | rs1260326 | - | - | T(C) | 0.009 (0.001) | <0.001 (0.009) |
| 10 | rs12956382 | rs11665601 | 0.90 | G(T) | -0.011 (0.001) | 0.001 (0.009) |
| 11 | rs133062 | - | - | A(G) | 0.009 (0.001) | 0.013 (0.009) |
| 12 | rs1513475 | - | - | C(T) | -0.010 (0.001) | -0.019 (0.010) |
| 13 | rs1542915 | - | - | C(A) | -0.008 (0.001) | 0.002 (0.009) |
| 14 | rs1568452 | - | - | T(C) | -0.012 (0.001) | 0.020 (0.009) |
| 15 | rs1594710 | - | - | T(G) | 0.007 (0.001) | -0.006 (0.009) |
| 16 | rs17598729 | - | - | C(T) | -0.011 (0.002) | -0.031 (0.012) |
| 17 | rs17731087 | - | - | T(C) | -0.016 (0.003) | 0.016 (0.019) |
| 18 | rs1997468 | - | - | T(C) | 0.011 (0.001) | 0.004 (0.009) |
| 19 | rs210600 | - | - | A(G) | 0.009 (0.002) | 0.010 (0.010) |
| 20 | rs2472297 | - | - | T(C) | -0.016 (0.002) | 0.011 (0.010) |
| 21 | rs2606222 | - | - | T(C) | 0.008 (0.001) | -0.010 (0.009) |
| 22 | rs2786529 | - | - | G(C) | -0.009 (0.002) | 0.002 (0.011) |
| 23 | rs2952896 | - | - | G(A) | -0.008 (0.001) | 0.006 (0.009) |
| 24 | rs4410790 | - | - | T(C) | 0.011 (0.001) | 0.001 (0.009) |
| 25 | rs480211 | - | - | C(T) | -0.008 (0.001) | -0.015 (0.009) |
| 26 | rs4957541 | - | - | G(T) | 0.008 (0.001) | 0.001 (0.009) |
| 27 | rs559411160 | N/A | N/A | - | - | - |
| 28 | rs61883187 | - | - | A(G) | 0.012 (0.002) | 0.025 (0.012) |
| 29 | rs61909866 | - | - | T(C) | -0.008 (0.001) | -0.018 (0.010) |
| 30 | rs6452788 | - | - | A(G) | -0.010 (0.002) | 0.029 (0.010) |
| 31 | rs66795913 | - | - | A(T) | -0.012 (0.001) | 0.011 (0.010) |
| 32 | rs76508707 | - | - | T(C) | 0.011 (0.001) | 0.007 (0.009) |
| 33 | rs7807990 | rs7808300 | 0.99 | C(A) | 0.008 (0.002) | 0.011 (0.010) |
| 34 | rs7947357 | - | - | A(G) | -0.008 (0.001) | -0.023 (0.009) |

| N | SNP | Proxy SNP | $r^2$ for proxy | Effect allele (reference) | β (SE) for hot drink temperature | β (SE) for MDD |
|---|---|---|---|---|---|---|
| 35 | rs835752 | - | - | A(G) | 0.008 (0.001) | -0.010 (0.010) |
| 36 | rs888405 | - | - | A(G) | -0.012 (0.002) | 0.024 (0.011) |
| 37 | rs9266224 | rs9266221 | 0.84 | T(C) | 0.009 (0.001) | 0.030 (0.010) |
| 38 | rs9271392 | rs9271550 | 0.97 | G(A) | -0.014 (0.002) | -0.020 (0.010) |
| 39 | rs9320913 | - | - | A(C) | -0.008 (0.001) | 0.013 (0.009) |
| 40 | rs9453572 | - | - | C(T) | -0.008 (0.001) | 0.005 (0.010) |

SNP, single nucleotide polymorphism; SE, standard error; MDD, major depressive disorder

Note: rs8019112 removed for being palindromic with intermediate allele frequencies. N/A indicates that proxy SNP with $r^2 > 0.8$ cannot be found.

#### Supplementary Table S20

Genome-wide significant SNPs for water intake

| N | SNP | Proxy SNP | $r^2$ for proxy | Effect allele (reference) | β (SE) for water intake | β (SE) for MDD |
|---|---|---|---|---|---|---|
| 1 | rs10145461 | - | - | T(G) | 0.012 (0.002) | -0.005 (0.009) |
| 2 | rs11012726 | - | - | C(T) | -0.013 (0.002) | -0.002 (0.010) |
| 3 | rs12753617 | N/A | N/A | - | - | - |
| 4 | rs1381273 | - | - | T(C) | 0.015 (0.002) | 0.015 (0.009) |
| 5 | rs1421085 | - | - | C(T) | 0.013 (0.002) | 0.016 (0.009) |
| 6 | rs150095288 | - | - | A(G) | -0.020 (0.003) | 0.009 (0.014) |
| 7 | rs17145750 | - | - | T(C) | -0.020 (0.003) | -0.015 (0.013) |
| 8 | rs2289292 | - | - | T(C) | -0.012 (0.002) | 0.015 (0.010) |
| 9 | rs2472297 | - | - | T(C) | -0.034 (0.002) | 0.011 (0.010) |
| 10 | rs28612302 | - | - | T(C) | 0.012 (0.002) | 0.014 (0.010) |
| 11 | rs34179251 | N/A | N/A | - | - | - |
| 12 | rs3933088 | - | - | C(T) | -0.012 (0.002) | -0.016 (0.009) |
| 13 | rs429358 | - | - | C(T) | 0.017 (0.003) | -0.040 (0.015) |
| 14 | rs4310286 | - | - | T(C) | 0.012 (0.002) | -0.016 (0.009) |
| 15 | rs4410790 | - | - | T(C) | 0.030 (0.002) | 0.001 (0.009) |
| 16 | rs4506023 | - | - | T(C) | -0.012 (0.002) | <0.001 (0.010) |
| 17 | rs4603502 | - | - | C(T) | -0.014 (0.002) | 0.004 (0.010) |
| 18 | rs4972509 | N/A | N/A | - | - | - |
| 19 | rs57545942 | - | - | T(C) | 0.020 (0.003) | -0.012 (0.013) |
| 20 | rs58242878 | rs60195615 | 0.96 | T(A) | -0.013 (0.002) | -0.011 (0.010) |
| 21 | rs58959216 | - | - | C(A) | -0.016 (0.003) | -0.006 (0.012) |
| 22 | rs7046160 | - | - | C(T) | 0.014 (0.002) | -0.025 (0.011) |
| 23 | rs7628633 | - | - | T(C) | -0.012 (0.002) | 0.009 (0.010) |
| 24 | rs7786041 | - | - | T(C) | 0.019 (0.003) | 0.002 (0.013) |
| 25 | rs779107 | - | - | C(A) | -0.012 (0.002) | -0.003 (0.009) |
| 26 | rs7938998 | - | - | C(A) | 0.015 (0.002) | 0.001 (0.009) |
| 27 | rs9419447 | - | - | A(C) | 0.011 (0.002) | -0.001 (0.009) |

SNP, single nucleotide polymorphism; SE, standard error; MDD, major depressive disorder

Notes: rs10758255 removed for being palindromic with intermediate allele frequencies. N/A indicates that proxy SNP with $r^2 > 0.8$ cannot be found.

#### Supplementary Table S21

Genome-wide significant SNPs for alcohol intake frequency

| N | SNP | Proxy SNP | $r^2$ for proxy | Effect allele (reference) | β (SE) for alcohol intake frequency | β (SE) for MDD |
|---|---|---|---|---|---|---|
| 1 | rs1004787 | - | - | G(A) | 0.023 (0.003) | 0.003 (0.009) |
| 2 | rs10278679 | - | - | G(A) | -0.020 (0.003) | 0.031 (0.009) |
| 3 | rs1078345 | - | - | G(A) | 0.028 (0.004) | 0.017 (0.011) |
| 4 | rs113011189 | N/A | N/A | - | - | - |
| 5 | rs117799466 | N/A | N/A | - | - | - |
| 6 | rs11787216 | - | - | T(C) | 0.026 (0.004) | -0.007 (0.010) |
| 7 | rs11940694 | - | - | A(G) | 0.043 (0.004) | -0.005 (0.009) |
| 8 | rs1229984 | - | - | T(C) | 0.281 (0.012) | 0.010 (0.026) |
| 9 | rs12967878 | - | - | C(T) | 0.027 (0.004) | -0.010 (0.011) |
| 10 | rs13014929 | - | - | C(G) | -0.025 (0.004) | -0.004 (0.012) |
| 11 | rs13102973 | - | - | T(C) | 0.020 (0.004) | 0.010 (0.009) |
| 12 | rs13135092 | - | - | G(A) | 0.049 (0.006) | -0.020 (0.019) |
| 13 | rs1421085 | - | - | C(T) | 0.021 (0.004) | 0.016 (0.009) |
| 14 | rs17690703 | rs112583797 | 0.83 | T(C) | 0.027 (0.004) | -0.005 (0.011) |
| 15 | rs1788030 | - | - | T(C) | 0.021 (0.003) | 0.020 (0.009) |
| 16 | rs202209188 | N/A | N/A | - | - | - |
| 17 | rs2111861 | - | - | G(A) | 0.020 (0.004) | 0.001 (0.009) |
| 18 | rs2244598 | - | - | T(C) | 0.020 (0.004) | -0.014 (0.010) |
| 19 | rs248558 | - | - | T(G) | 0.022 (0.003) | 0.024 (0.009) |
| 20 | rs2717063 | - | - | C(A) | 0.019 (0.004) | -0.010 (0.009) |
| 21 | rs33705 | - | - | T(C) | -0.025 (0.004) | 0.024 (0.009) |
| 22 | rs35105141 | - | - | T(C) | 0.025 (0.004) | -0.011 (0.009) |
| 23 | rs4239694 | - | - | G(T) | -0.022 (0.004) | <0.001 (0.010) |
| 24 | rs4417025 | - | - | A(G) | -0.020 (0.004) | 0.009 (0.010) |
| 25 | rs4726481 | - | - | T(G) | 0.022 (0.004) | 0.001 (0.010) |
| 26 | rs4800487 | - | - | G(A) | -0.031 (0.003) | 0.017 (0.009) |
| 27 | rs489062 | - | - | A(G) | 0.019 (0.003) | -0.016 (0.009) |
| 28 | rs4970394 | N/A | N/A | - | - | - |
| 29 | rs56119718 | rs2622099 | 0.99 | C(T) | -0.020 (0.004) | -0.017 (0.010) |
| 30 | rs56189237 | - | - | A(G) | 0.029 (0.005) | -0.011 (0.013) |
| 31 | rs56228311 | - | - | T(G) | -0.020 (0.004) | -0.002 (0.010) |
| 32 | rs57281063 | - | - | A(G) | -0.028 (0.003) | 0.004 (0.009) |
| 33 | rs61873510 | - | - | T(G) | 0.023 (0.004) | 0.015 (0.010) |
| 34 | rs62305780 | - | - | G(C) | -0.053 (0.006) | -0.003 (0.015) |

| N | SNP | Proxy SNP | $r^2$ for proxy | Effect allele (reference) | β (SE) for alcohol intake frequency | β (SE) for MDD |
|---|---|---|---|---|---|---|
| 35 | rs62339673 | - | - | C(A) | -0.020 (0.004) | 0.002 (0.010) |
| 36 | rs62466318 | - | - | T(C) | -0.026 (0.004) | -0.015 (0.012) |
| 37 | rs650558 | - | - | T(C) | 0.022 (0.004) | -0.010 (0.011) |
| 38 | rs7117115 | rs7935344 | 0.96 | G(A) | -0.025 (0.004) | 0.012 (0.009) |
| 39 | rs7196706 | - | - | T(C) | 0.023 (0.004) | -0.009 (0.010) |
| 40 | rs72806111 | - | - | G(A) | -0.022 (0.004) | 0.012 (0.011) |
| 41 | rs73050128 | - | - | A(C) | -0.026 (0.005) | -0.032 (0.012) |
| 42 | rs73193340 | - | - | G(A) | 0.035 (0.005) | 0.017 (0.013) |
| 43 | rs7460106 | N/A | N/A | - | - | - |
| 44 | rs748919 | - | - | C(T) | 0.025 (0.004) | -0.012 (0.011) |
| 45 | rs780094 | - | - | T(C) | 0.049 (0.004) | -0.001 (0.009) |
| 46 | rs780569 | - | - | T(A) | -0.022 (0.004) | 0.007 (0.010) |
| 47 | rs7991335 | - | - | C(A) | 0.023 (0.004) | -0.002 (0.010) |
| 48 | rs80292319 | - | - | C(T) | -0.041 (0.007) | -0.023 (0.018) |
| 49 | rs8049739 | - | - | G(C) | 0.034 (0.004) | -0.007 (0.009) |
| 50 | rs838145 | - | - | G(A) | -0.023 (0.003) | 0.009 (0.009) |
| 51 | rs8614 | - | - | A(C) | 0.027 (0.004) | 0.001 (0.012) |
| 52 | rs9349379 | - | - | G(A) | -0.020 (0.003) | 0.005 (0.009) |
| 53 | rs9372625 | - | - | A(G) | -0.026 (0.004) | 0.004 (0.009) |
| 54 | rs9814516 | - | - | T(G) | -0.026 (0.004) | -0.004 (0.010) |
| 55 | rs9842406 | - | - | G(T) | -0.025 (0.003) | -0.006 (0.009) |
| 56 | rs9891943 | - | - | C(T) | 0.025 (0.004) | 0.021 (0.011) |
| 57 | rs9923768 | - | - | G(A) | 0.020 (0.004) | 0.002 (0.009) |

SNP, single nucleotide polymorphism; SE, standard error; MDD, major depressive disorder

Notes: rs1104608 and rs13097366 removed for being palindromic with intermediate allele frequencies. N/A indicates that proxy SNP with $r^2 > 0.8$ cannot be found.

#### Supplementary Table S22

Genome-wide reported SNPs for major depressive disorder ($P< 1 \times 10^{-6}$)

| N | SNP | Proxy SNP | $r^2$ for proxy | Effect allele (reference) | β (SE) for MDD | β (SE) for cooked vegetable intake | β (SE) for salad / raw vegetable intake | β (SE) for fresh fruit intake | β (SE) for dried fruit intake | β (SE) for oily fish intake |
|---|---|---|---|---|---|---|---|---|---|---|
| 1 | rs1081458 | - | - | A(T) | 0.060 (0.012) | <0.001 (0.002) | 0.009 (0.002) | 0.003 (0.002) | 0.004 (0.003) | 0.002 (0.003) |
| 2 | rs10825942 | - | - | T(G) | -0.048 (0.009) | -0.002 (0.002) | <0.001 (0.002) | <0.001 (0.001) | <0.001 (0.002) | 0.001 (0.002) |
| 3 | rs1256112 | - | - | T(C) | 0.044 (0.009) | -0.001 (0.002) | -0.005 (0.002) | -0.001 (0.001) | -0.002 (0.002) | -0.004 (0.002) |
| 4 | rs17499892 | - | - | A(C) | -0.050 (0.009) | <0.001 (0.002) | 0.001 (0.002) | -0.002 (0.001) | -0.004 (0.002) | 0.002 (0.002) |
| 5 | rs1936365 | - | - | C(G) | -0.069 (0.013) | -0.004 (0.002) | -0.010 (0.002) | -0.001 (0.002) | -0.010 (0.003) | -0.017 (0.003) |
| 6 | rs1950829[a] | - | - | A(G) | 0.054 (0.009) | 0.001 (0.002) | 0.001 (0.002) | -0.004 (0.001) | -0.002 (0.002) | -0.001 (0.002) |
| 7 | rs2012697 | - | - | T(C) | -0.048 (0.009) | -0.004 (0.002) | -0.006 (0.002) | -0.010 (0.001) | -0.014 (0.002) | -0.011 (0.002) |
| 8 | rs2060886 | - | - | T(C) | -0.046 (0.009) | <0.001 (0.002) | -0.001 (0.002) | -0.002 (0.001) | <0.001 (0.002) | -0.002 (0.002) |
| 9 | rs2451828 | - | - | T(C) | 0.159 (0.031) | <0.001 (0.006) | -0.002 (0.006) | 0.008 (0.005) | 0.003 (0.006) | 0.017 (0.007) |
| 10 | rs281265 | - | - | A(C) | -0.046 (0.009) | 0.004 (0.002) | <0.001 (0.002) | 0.004 (0.001) | 0.008 (0.002) | 0.009 (0.002) |
| 11 | rs34382743 | - | - | T(C) | 0.046 (0.009) | 0.001 (0.002) | -0.002 (0.002) | -0.002 (0.001) | -0.002 (0.002) | -0.001 (0.002) |
| 12 | rs4776768 | - | - | T(C) | -0.050 (0.010) | -0.003 (0.002) | -0.004 (0.002) | <0.001 (0.002) | 0.001 (0.002) | <0.001 (0.002) |
| 13 | rs4811079 | - | - | C(G) | 0.049 (0.009) | 0.002 (0.002) | -0.001 (0.002) | <0.001 (0.001) | -0.002 (0.002) | 0.003 (0.002) |
| 14 | rs586275 | - | - | A(G) | -0.059 (0.012) | -0.002 (0.002) | <0.001 (0.002) | -0.001 (0.002) | 0.001 (0.002) | -0.004 (0.003) |
| 15 | rs6689226 | - | - | T(C) | 0.050 (0.009) | -0.003 (0.002) | <0.001 (0.002) | 0.001 (0.001) | -0.001 (0.002) | 0.005 (0.002) |
| 16 | rs6832890 | - | - | C(G) | -0.060 (0.011) | -0.006 (0.002) | -0.005 (0.002) | -0.002 (0.002) | -0.005 (0.002) | -0.005 (0.003) |
| 17 | rs76025409[a] | - | - | C(G) | 0.057 (0.010) | 0.002 (0.002) | <0.001 (0.002) | -0.002 (0.001) | -0.001 (0.002) | -0.004 (0.002) |
| 18 | rs78676209 | - | - | C(G) | -0.104 (0.020) | 0.002 (0.004) | 0.005 (0.004) | 0.002 (0.003) | 0.001 (0.004) | 0.006 (0.005) |

SNP, single nucleotide polymorphism; SE, standard error; MDD, major depressive disorder;

[a]: p-value for genome-wide association < 5 × $10^{-8}$

#### Supplementary Table S22

Genome-wide reported SNPs for major depressive disorder ($P< 1 \times 10^{-6}$) (continued)

| N | SNP | β (SE) for non-oily fish intake | β (SE) for processed meat intake | β (SE) for poultry intake | β (SE) for beef intake | β (SE) for lamb/mutton intake | β (SE) for pork intake | β (SE) for cheese intake | β (SE) for bread intake |
|---|---|---|---|---|---|---|---|---|---|
| 1 | rs1081458 | 0.007 (0.002) | 0.007 (0.003) | -0.006 (0.003) | 0.002 (0.003) | -0.003 (0.002) | -0.002 (0.002) | 0.004 (0.003) | 0.005 (0.003) |
| 2 | rs10825942 | 0.001 (0.002) | -0.003 (0.002) | -0.001 (0.002) | -0.004 (0.002) | 0.001 (0.002) | <0.001 (0.002) | -0.002 (0.003) | -0.001 (0.002) |
| 3 | rs1256112 | -0.002 (0.002) | -0.001 (0.002) | <0.001 (0.002) | -0.003 (0.002) | -0.001 (0.002) | -0.001 (0.002) | 0.003 (0.003) | 0.002 (0.002) |
| 4 | rs17499892 | <0.001 (0.002) | -0.002 (0.002) | 0.004 (0.002) | 0.002 (0.002) | 0.002 (0.002) | 0.003 (0.002) | <0.001 (0.003) | 0.001 (0.002) |
| 5 | rs1936365 | -0.008 (0.002) | 0.002 (0.003) | 0.005 (0.003) | -0.001 (0.003) | <0.001 (0.002) | 0.002 (0.002) | 0.005 (0.003) | 0.012 (0.003) |
| 6 | rs1950829[a] | 0.001 (0.002) | 0.002 (0.002) | -0.002 (0.002) | <0.001 (0.002) | -0.002 (0.002) | -0.001 (0.002) | -0.002 (0.003) | 0.001 (0.002) |
| 7 | rs2012697 | -0.005 (0.002) | 0.014 (0.002) | 0.001 (0.002) | 0.014 (0.002) | 0.008 (0.002) | 0.006 (0.002) | <0.001 (0.003) | 0.006 (0.002) |
| 8 | rs2060886 | -0.001 (0.002) | 0.005 (0.002) | 0.003 (0.002) | 0.005 (0.002) | 0.001 (0.002) | 0.003 (0.002) | 0.003 (0.003) | 0.005 (0.002) |
| 9 | rs2451828 | 0.009 (0.006) | 0.006 (0.008) | -0.001 (0.007) | <0.001 (0.007) | 0.003 (0.006) | -0.005 (0.006) | -0.006 (0.009) | -0.008 (0.008) |
| 10 | rs281265 | 0.001 (0.002) | 0.003 (0.002) | -0.001 (0.002) | 0.002 (0.002) | 0.001 (0.002) | <0.001 (0.002) | 0.001 (0.003) | 0.003 (0.002) |
| 11 | rs34382743 | 0.002 (0.002) | -0.004 (0.002) | 0.002 (0.002) | 0.004 (0.002) | 0.007 (0.002) | -0.001 (0.002) | -0.002 (0.003) | -0.002 (0.002) |
| 12 | rs4776768 | <0.001 (0.002) | 0.003 (0.003) | -0.003 (0.002) | <0.001 (0.002) | 0.001 (0.002) | -0.002 (0.002) | 0.002 (0.003) | 0.001 (0.003) |
| 13 | rs4811079 | 0.003 (0.002) | 0.003 (0.002) | 0.005 (0.002) | 0.002 (0.002) | 0.003 (0.002) | 0.005 (0.002) | -0.003 (0.003) | -0.003 (0.002) |
| 14 | rs586275 | -0.001 (0.002) | <0.001 (0.003) | 0.001 (0.003) | -0.003 (0.002) | -0.003 (0.002) | -0.001 (0.002) | 0.002 (0.003) | -0.001 (0.003) |
| 15 | rs6689226 | 0.007 (0.002) | 0.002 (0.002) | <0.001 (0.002) | -0.004 (0.002) | -0.004 (0.002) | -0.003 (0.002) | -0.007 (0.003) | 0.001 (0.002) |
| 16 | rs6832890 | -0.001 (0.002) | 0.004 (0.003) | 0.001 (0.003) | 0.003 (0.002) | <0.001 (0.002) | 0.003 (0.002) | -0.006 (0.003) | -0.004 (0.003) |
| 17 | rs76025409[a] | -0.001 (0.002) | 0.004 (0.002) | <0.001 (0.002) | 0.002 (0.002) | 0.001 (0.002) | 0.001 (0.002) | -0.002 (0.003) | 0.001 (0.002) |
| 18 | rs78676209 | 0.004 (0.004) | -0.008 (0.006) | -0.006 (0.005) | -0.002 (0.005) | -0.005 (0.004) | -0.008 (0.004) | -0.005 (0.006) | -0.007 (0.005) |

SNP, single nucleotide polymorphism; SE, standard error; MDD, major depressive disorder;

[a]: p-value for genome-wide association < $5 \times 10^{-8}$

**Supplementary Table S22**

Genome-wide reported SNPs for major depressive disorder ($P< 1 \times 10^{-6}$) (continued)

| N | SNP | β (SE) for cereal intake | β (SE) for salt added to food | β (SE) for tea intake | β (SE) for coffee intake | β (SE) for hot drink temperature | β (SE) for water intake | β (SE) for alcohol intake frequency |
|---|---|---|---|---|---|---|---|---|
| 1 | rs1081458 | 0.001 (0.003) | 0.002 (0.003) | -0.007 (0.003) | <0.001 (0.002) | -0.003 (0.002) | 0.003 (0.003) | -0.001 (0.005) |
| 2 | rs10825942 | 0.001 (0.002) | -0.002 (0.002) | <0.001 (0.002) | -0.004 (0.002) | 0.001 (0.001) | 0.002 (0.002) | 0.002 (0.003) |
| 3 | rs1256112 | 0.001 (0.002) | -0.004 (0.002) | 0.004 (0.002) | 0.002 (0.002) | -0.004 (0.001) | -0.001 (0.002) | -0.005 (0.003) |
| 4 | rs17499892 | -0.002 (0.002) | 0.005 (0.002) | -0.005 (0.002) | 0.001 (0.002) | -0.003 (0.001) | -0.006 (0.002) | -0.004 (0.004) |
| 5 | rs1936365 | -0.001 (0.003) | -0.005 (0.003) | 0.003 (0.003) | <0.001 (0.002) | -0.008 (0.002) | -0.006 (0.003) | 0.009 (0.005) |
| 6 | rs1950829[a] | 0.001 (0.002) | 0.005 (0.002) | 0.006 (0.002) | <0.001 (0.002) | 0.001 (0.001) | <0.001 (0.002) | 0.004 (0.003) |
| 7 | rs2012697 | -0.006 (0.002) | 0.006 (0.002) | -0.005 (0.002) | -0.003 (0.002) | 0.001 (0.001) | -0.010 (0.002) | -0.010 (0.003) |
| 8 | rs2060886 | <0.001 (0.002) | 0.002 (0.002) | -0.005 (0.002) | 0.001 (0.002) | -0.001 (0.001) | <0.001 (0.002) | -0.014 (0.003) |
| 9 | rs2451828 | -0.007 (0.007) | 0.003 (0.007) | -0.017 (0.008) | -0.001 (0.006) | -0.002 (0.005) | -0.002 (0.007) | -0.019 (0.012) |
| 10 | rs281265 | <0.001 (0.002) | 0.004 (0.002) | 0.004 (0.003) | -0.004 (0.002) | 0.001 (0.001) | <0.001 (0.002) | 0.007 (0.004) |
| 11 | rs34382743 | <0.001 (0.002) | -0.010 (0.002) | <0.001 (0.002) | 0.003 (0.002) | -0.003 (0.001) | 0.003 (0.002) | 0.001 (0.004) |
| 12 | rs4776768 | -0.001 (0.002) | 0.001 (0.002) | -0.002 (0.003) | -0.001 (0.002) | <0.001 (0.001) | -0.001 (0.002) | 0.001 (0.004) |
| 13 | rs4811079 | -0.004 (0.002) | <0.001 (0.002) | 0.005 (0.002) | -0.002 (0.002) | 0.003 (0.001) | <0.001 (0.002) | 0.003 (0.004) |
| 14 | rs586275 | <0.001 (0.002) | -0.002 (0.002) | -0.002 (0.003) | 0.001 (0.002) | -0.002 (0.002) | -0.002 (0.003) | -0.009 (0.004) |
| 15 | rs6689226 | -0.002 (0.002) | -0.002 (0.002) | -0.001 (0.002) | 0.003 (0.002) | 0.002 (0.001) | <0.001 (0.002) | 0.002 (0.004) |
| 16 | rs6832890 | -0.007 (0.003) | 0.008 (0.003) | -0.006 (0.003) | <0.001 (0.002) | -0.002 (0.002) | 0.002 (0.003) | -0.007 (0.004) |
| 17 | rs76025409[a] | -0.006 (0.002) | 0.013 (0.002) | -0.011 (0.003) | -0.001 (0.002) | -0.001 (0.001) | 0.006 (0.002) | 0.008 (0.004) |
| 18 | rs78676209 | -0.002 (0.005) | -0.012 (0.005) | 0.003 (0.006) | 0.002 (0.004) | -0.008 (0.003) | 0.001 (0.005) | -0.002 (0.008) |

SNP, single nucleotide polymorphism; SE, standard error; MDD, major depressive disorder;

[a]: p-value for genome-wide association $< 5 \times 10^{-8}$

#### Supplementary Table S23

Tests for detecting horizontal pleiotropy

| Exposure | Outcome | MR-PRESSO global test | | Heterogeneity Cochrane's Q | | | Heterogeneity MR-Egger | | | Intercept test in MR-Egger regression | | |
|---|---|---|---|---|---|---|---|---|---|---|---|---|
| | | RSS | p-value | Q | df | p-value | Q | df | p-value | intercept | se | p-value |
| Non-oily fish intake | MDD | 4.59 | 0.679 | 2.73 | 5 | 0.742 | 2.67 | 4 | 0.614 | 0.005 | 0.02 | 0.824 |
| Beef intake [a] | MDD | 68.61 | <0.001 | 54.40 | 8 | <0.001 | 53.64 | 7 | <0.001 | 0.026 | 0.08 | 0.762 |
| Cereal intake | MDD | 22.16 | 0.466 | 20.13 | 20 | 0.450 | 19.93 | 19 | 0.399 | 0.004 | 0.01 | 0.668 |

MR, Mendelian randomization; RSS, residual sum of square; df, degree of freedom; se, standard error; MDD, major depressive disorder;

[a]: MR-Egger test (Q = 7.44; p-value = 0.114) and Cochran's Q test (Q = 7.93; p-value = 0.160) for heterogeneity were no longer statistically significant after correcting for horizontal pleiotropy via outlier removal.

### Supplementary Figure Legends

##### Supplementary Fig. S1

Mendelian randomization results for estimating the causal effects of non-oily fish intake on major depressive disorder. (A) Scatter plot showing the effects of SNPs on non-oily fish intake versus major depressive disorder. (B) Forest plot of Mendelian randomization effect size for non-oily fish intake on major depressive disorder.

##### Supplementary Fig. S2

Mendelian randomization results for estimating the causal effects of cereal intake on major depressive disorder. (A) Scatter plot showing the effects of SNPs on cereal intake versus major depressive disorder. (B) Forest plot of Mendelian randomization effect size for cereal intake on major depressive disorder.

##### Supplementary Fig. S3

Funnel plot of the effect of beef intake on major depressive disorder.

##### Supplementary Fig. S4

Funnel plot of the effect of non-oily fish intake on major depressive disorder.

**Supplementary Fig. S5**

Funnel plot of the effect of cereal intake on major depressive disorder.

**Supplementary Fig. S6**

Leave-one-out analysis of the effect of non-oily fish intake on major depressive disorder.

**Supplementary Fig. S7**

Leave-one-out analysis of the effect of cereal intake on major depressive disorder.

**Supplementary Fig. S1**

Mendelian randomization results for estimating the causal effects of non-oily fish intake on major depressive disorder. (**A**) Scatter plot showing the effects of SNPs on non-oily fish intake versus major depressive disorder. (**B**) Forest plot of Mendelian randomization effect size of non-oily fish intake on major depressive disorder.

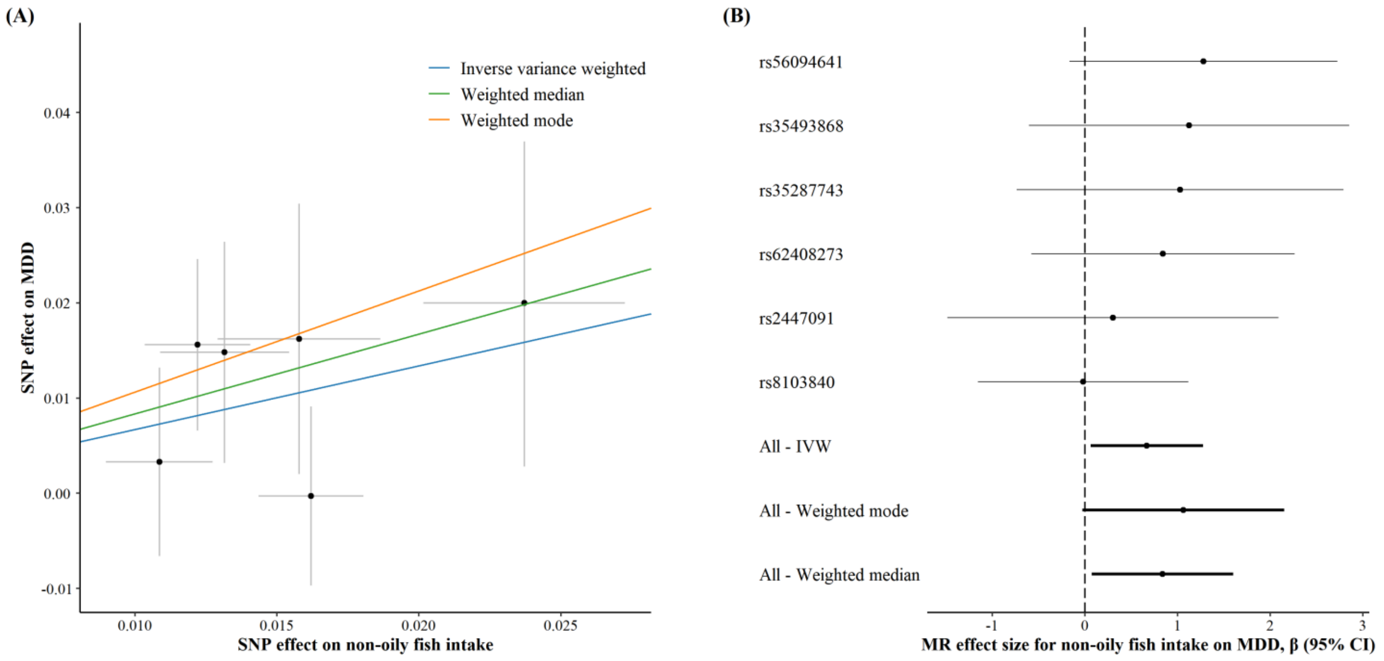

#### Supplementary Fig. S2

Mendelian randomization results for estimating the causal effects of cereal intake on major depressive disorder. (**A**) Scatter plot showing the effects of SNPs on cereal intake versus major depressive disorder. (**B**) Forest plot of Mendelian randomization effect size of cereal intake on major depressive disorder.

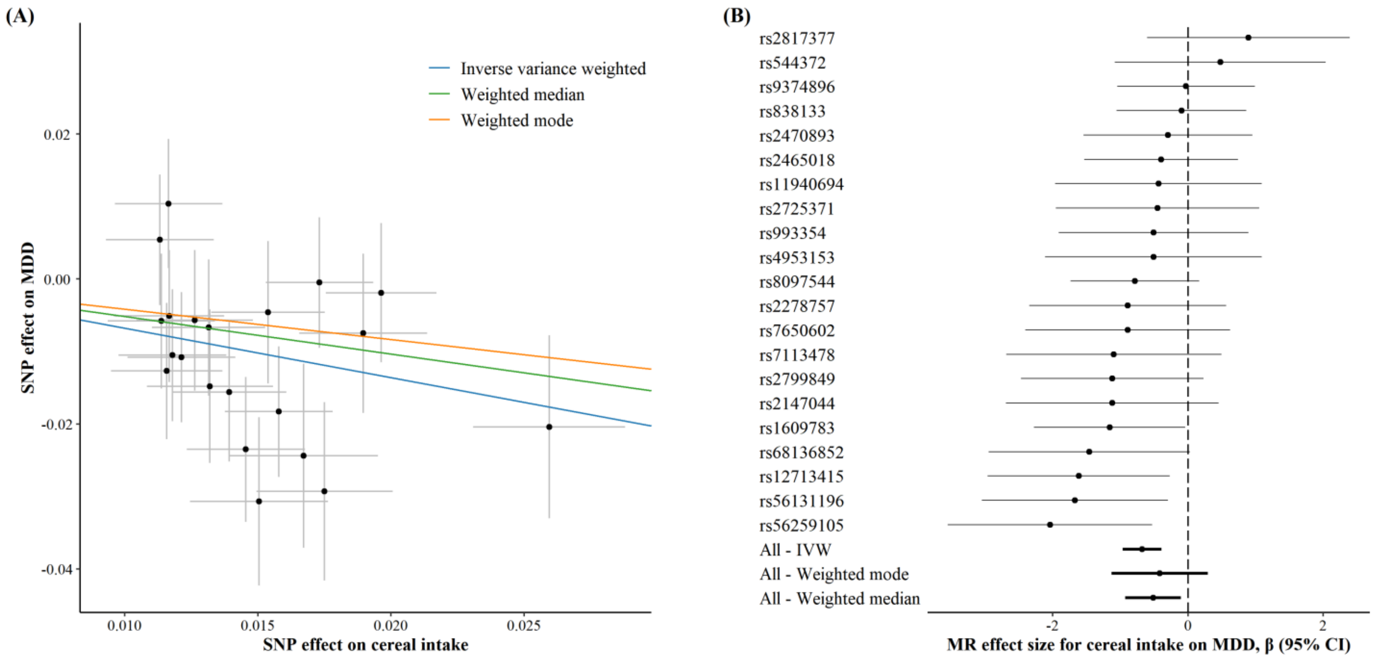

**Supplementary Fig. S3**

Funnel plot of the effect of beef intake on major depressive disorder.

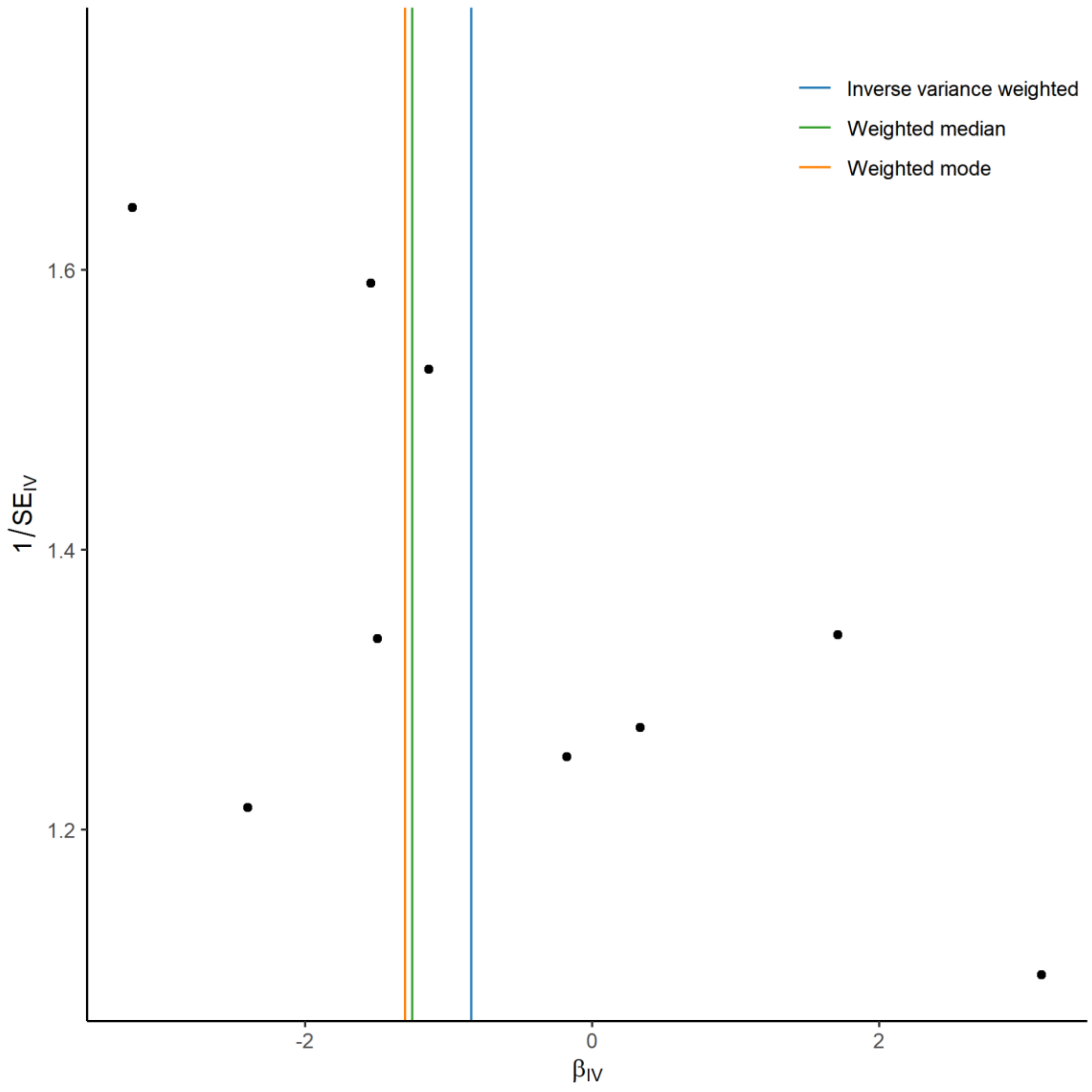

**Supplementary Fig. S4**

Funnel plot of the effect of non-oily fish intake on major depressive disorder.

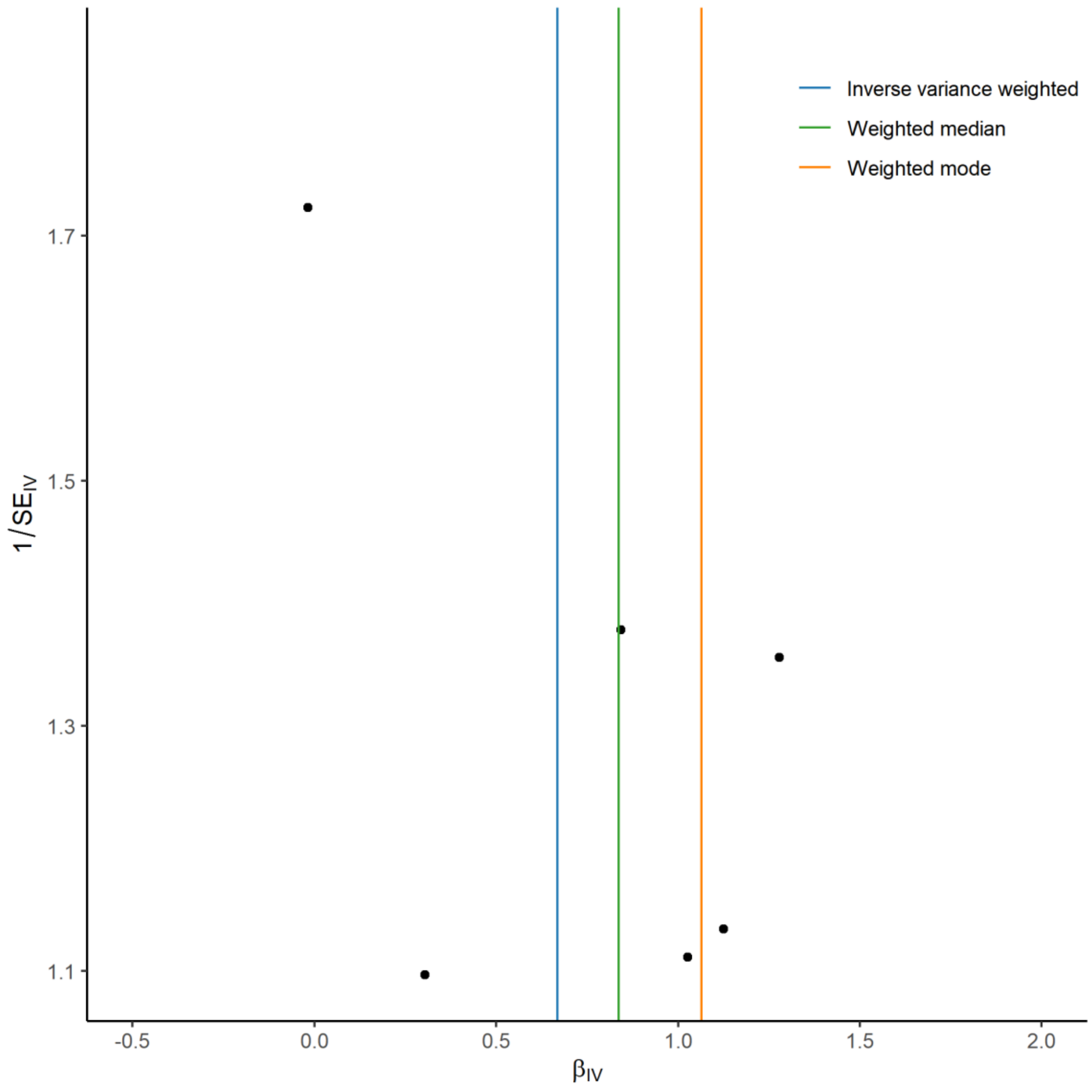

**Supplementary Fig. S5**

Funnel plot of the effect of cereal intake on major depressive disorder.

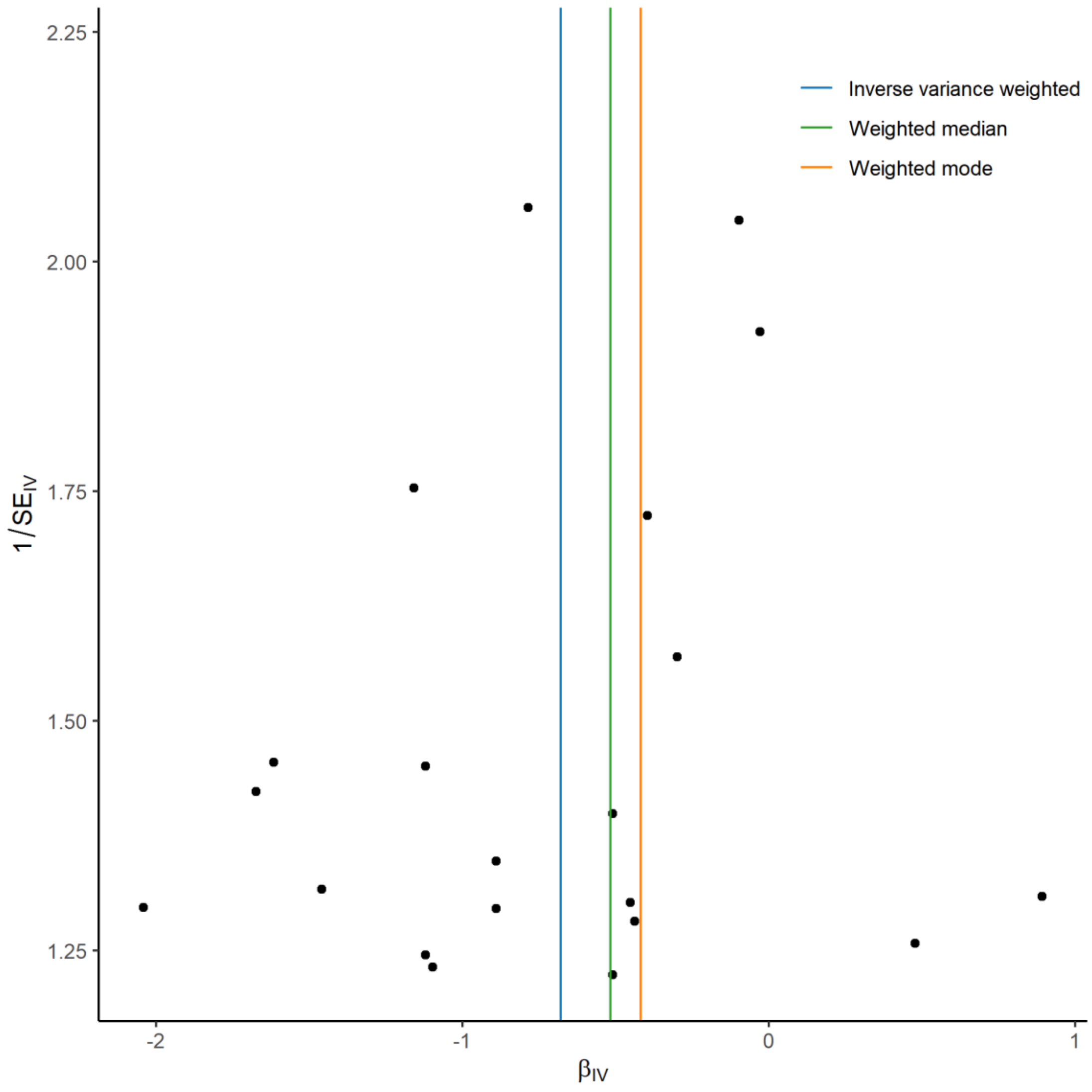

**Supplementary Fig. S6**

Leave-one-out analysis of the effect of non-oily fish intake on major depressive disorder.

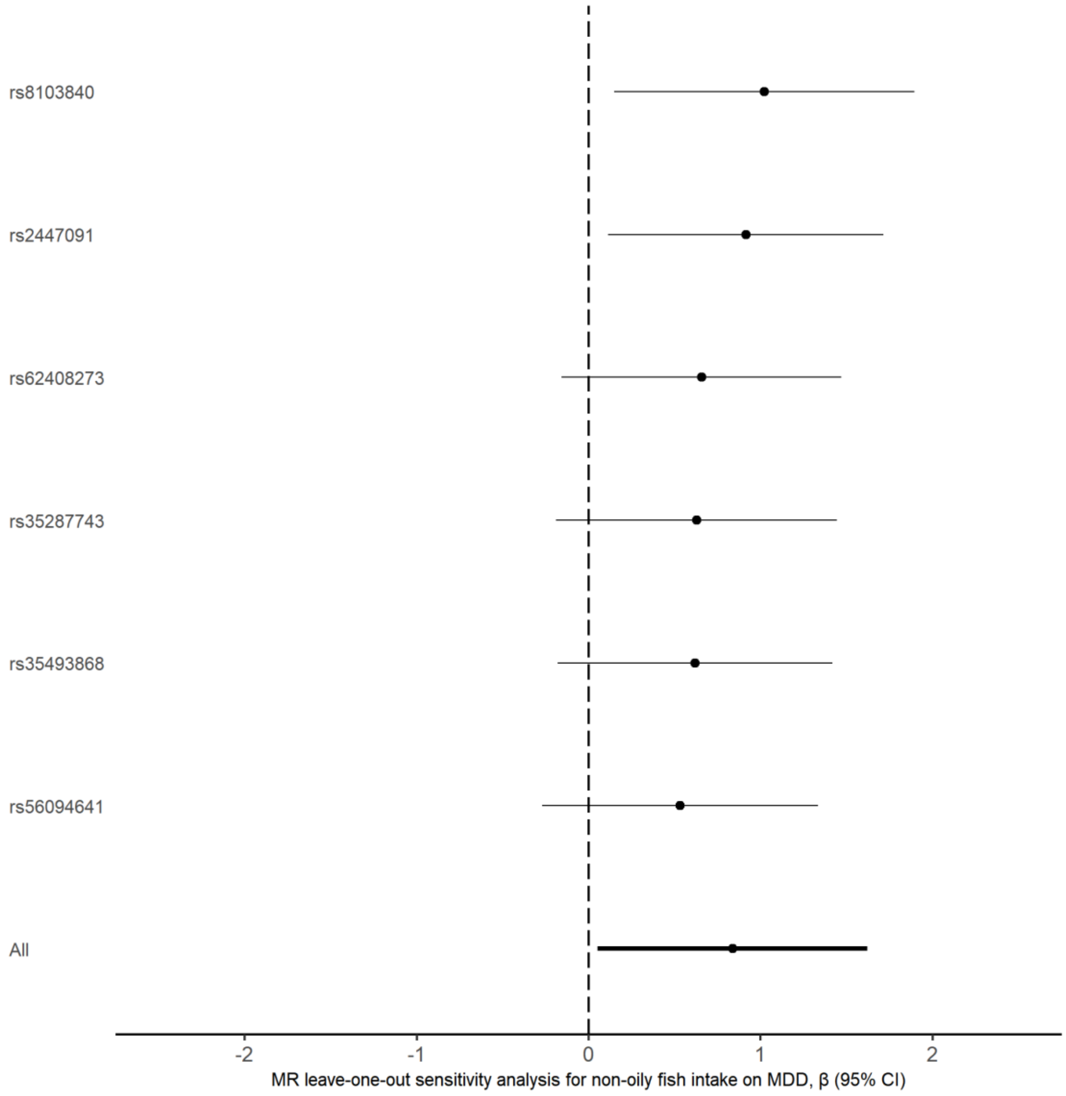

#### Supplementary Fig. S7

Leave-one-out analysis of the effect of cereal intake on major depressive disorder.

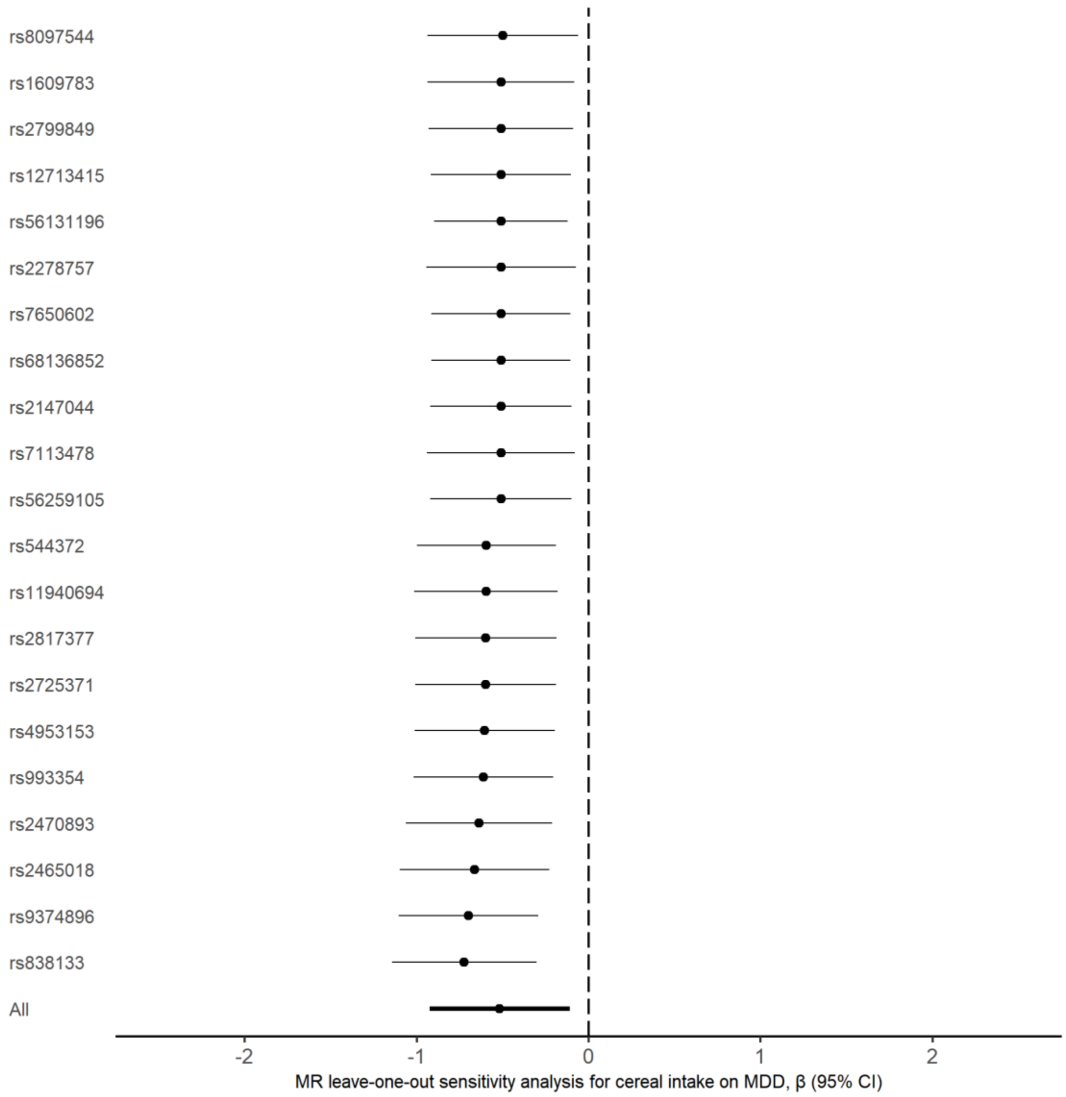
